# Research Transparency in 59 Disciplines of Clinical Medicine: A Meta-Research Study

**DOI:** 10.1101/2024.04.08.24305416

**Authors:** Ahmad Sofi-Mahmudi, Eero Raittio, Sergio E. Uribe, Sahar Khademioore, Dena Zeraatkar, Lawrence Mbuagbaw, Lex M. Bouter, Karen A. Robinson

## Abstract

**Background:** Transparency in health research is crucial as it allows for the scrutiny and replication of findings, fosters confidence in scientific outcomes, and ultimately contributes to the advancement of knowledge and the betterment of society.

**Aim:** We aimed to assess five transparency practices in scientific publications (data availability, code availability, protocol registration, conflicts of interest (COI) and funding disclosures) from open-access articles published in medical journals.

**Methods:** We searched and exported all open-access articles from Science Citation Index Expanded (SCIE)-indexed journals through the Europe PubMed Central database published until March 16, 2024. Basic journal- and article-related information was retrieved from the database. We then assessed five transparency practices in the articles using the *rtransparent* package in R.

**Results:** The analysis included 2,002,955 open-access articles from SCIE-indexed medical journals (open-access percentage=59.0%). Of these, 87.5% (95% CI: 87.4%-87.5%) disclosed COI and 80.1% (95% CI: 80.0%-80.1%) disclosed funding. Protocol registration was declared in 6.6% (95% CI: 6.6%-6.6%), data sharing in 7.6% (95% CI: 7.6%-7.6%), and code sharing in 1.4% (95% CI: 1.4%-1.4%) of the articles. More than 76.0% declared at least two transparency practices, while all five practices were declared in less than 0.02%. The data showed an increasing trend in all transparency practices since the late 2000s. Articles published in journals with higher impact factors and articles receiving more citations had increased odds of COI and funding disclosures, as well as data and code sharing. There were notable differences in transparency practices across the disciplines.

**Conclusion:** While most articles had COI and funding disclosures, adherence to other transparency practices was grossly insufficient. To increase protocol registration, data, and code sharing, much stronger incentives and mandates are needed from all stakeholders.

## Background

Recent recognition of health research transparency, essential for accountability, has resulted in stringent disclosure requirements by academic, medical institutions, and voluntary industry and publisher policies (1,2). Although global regulatory bodies, funding agencies, and ethics boards supervise medical research, transparency and disclosure practices are still inconsistent and incomplete (3,4). Research conducted by industry, academia, or their collaboration equally has deficiencies in transparency (2,5). Despite publishers’ policies, ethical mandates, and mission statements (6), academic medical centers show poor performance and significant variation in disseminating clinical trial results following transparency practices (7). Transparency in research is pivotal for accountability and trust in results and upholds the ethical responsibilities of researchers, editors, publishers, and funders (8–12). Inefficient use of primarily public or non-profit research funding significantly disadvantages patients and society. Therefore, it is crucial to continuously improve, investigate and monitor research transparency. Key indicators of research transparency include data sharing, code sharing, disclosures of conflicts of interest, funding acknowledgments, and protocol registration (13). Data sharing is more common in non-COVID-19 articles (12%) than in COVID-19 studies (4%) (14). A systematic review of 105 meta-research studies, analyzing 2,121,580 articles across 31 specialties, uncovered substantial transparency challenges (15). Issues include low declared (8%) and actual (2%) public data availability, minimal public code sharing (<0.5%), and inconsistent journal data-sharing policy adherence. This review also noted discrepancies between declared and actual data sharing practices and challenges in privately obtaining data and code from authors (15). These findings highlight the pressing need to enhance transparency, particularly during public health crises.

Our meta-research study assessed research transparency regarding data and code sharing, conflict of interest (COI) and funding disclosure, and protocol registration across all medical specialties. Through programmatic and comprehensive analysis, we aimed to identify patterns and areas most needing improvement.

## Methods

The protocol of this descriptive study was published on the Open Science Framework (OSF) website (https://doi.org/10.17605/OSF.IO/J57BG). All the code and data associated with the study were shared through both its OSF repository (https://osf.io/zbc6p/) and GitHub (https://github.com/choxos/medical-transparency). To ensure transparency and facilitate the reproducibility of our analyses, a PDF document containing the codes and corresponding outputs is provided in Appendix 1.

### Data sources and study selection

Initially, we searched records within journals listed in the 59 disciplines of the “Clinical Medicine” section in the Science Citation Index Expanded (SCIE) version 2020. This search was performed using the Europe PubMed Central (EPMC) database until February 28, 2022. We updated the search on March 16, 2024. The EPMC database encompasses all records found in PubMed and PubMed Central records and allows automated retrieval of full-texts of EPMC open-access records. Remarkably, what is being called here “EPMC open-access articles” do not include all “open-access” labeled articles (e.g., by the journals/publishers), because some of those articles are still subject to traditional copyright restrictions. Thus, their full texts cannot be accessed via EPMC.

Using the *metareadr* package (16), we retrieved the full texts of all identified open-access records in XML format from the EPMC database. Concurrently, we extracted descriptive details for each journal and article, including publisher, publication year, and citations linked to the article and journal, directly from the EPMC database. We only included research articles (`PUB_TYPE:”research-article”`) and reviews (`PUB_TYPE:”review-article” OR PUB_TYPE:”systematic-review”`) for our final analyses.

To identify trials, we created a database of trials by searching for articles that had `(PUB_TYPE:”Randomized Controlled Trial” OR PUB_TYPE:”Clinical Trial” OR PUB_TYPE:”Clinical Trial, Phase III” OR PUB_TYPE:”Clinical Trial, Phase IV”)` tags as their article type using the *europepmc* package. Articles in our data set that appeared in the trials database were tagged as trials. Similarly, articles tagged as `(PUB_TYPE:”review-article” OR PUB_TYPE:”systematic-review”)` were detected as reviews.

We used the Scimago Journal & Country Rank 2022 (SJR, https://www.scimagojr.com) to extract the publisher of the journals.

### Data extraction and synthesis

We used the *rtransparent* package (17), a validated and automated programmatic tool (13), to identify five transparent practices from the full texts we were able to download from EPMC:

1. *Data sharing*: The accessibility of data or metadata obtained during a research study, typically through public repositories or inclusion as supplementary materials accompanying the published work.
2. *Code sharing:* The disclosure of computer code or scripts employed for data analysis in research facilitates the replication of results and enables the broader utilization of the study’s methodologies.
3. *Conflict of interest (COI) disclosure*s: The public acknowledgment of potential conflicts of interest that could impact the research, commonly presented within a designated publication section.
4. *Funding disclosures*: The disclosure of the sources of financial support for the research, promoting transparency regarding the possible influence of funding organizations.
5. *Protocol registration:* The public disclosure of research protocols before conducting a study designed to reduce bias and increase transparency in the research process.

The package uses a standardized vocabulary to identify transparency indicators in EPMC XML files. It detects keywords related to COI disclosure, such as “conflicts of interest,” “competing interests,” or “nothing to disclose,” in article section titles or bodies. The tool recognizes all mentions of COI and funding disclosures and treats “nothing to disclose” statements as an indication of transparency, similar to actual conflict disclosures.

To assess data and code sharing, the *rtransparent* tool detects mentions that indicate that they are shared either as supplemental content, in general repositories (e.g., figshare, OSF, GitHub), or in field-specific repositories (e.g., dbSNP, ProteomeXchange, GenomeRNAi). Items that state “data available upon request” are not considered data sharing due to the unlikelihood of data acquisition (18). The *rtransparent* tool demonstrates robust validation, with high sensitivity and specificity for detecting transparency indicators: conflict of interest disclosure (sensitivity 99.2%, specificity 99.5%), funding disclosure (sensitivity 99.7%, specificity 98.1%), protocol registration (sensitivity 95.5%, specificity 99.7%), data sharing (sensitivity 75.8%, specificity 98.6%), and code sharing (sensitivity 58.7%, specificity 99.7%) (13).

### Data analysis

First, we computed the percentage of articles with full-texts available via EPMC (EPMC open-access records) out of the total number of articles within the database. Alongside providing descriptive statistics for the obtained sample, we reported transparency practices categorized by publication type. We also determined and reported the number of transparency practices that articles with available full-text declared within each publication type, ranging from 0 to 5 practices. Furthermore, we charted differences in transparency practices between 59 distinct clinical medicine disciplines over time. The sensitivity and specificity of the *rtransparent* tool (17) were used to generate 95% confidence intervals (CIs) for the estimates of the transparency practices. We used visual presentation and Pearson’s product-moment correlation to analyze the yearly trend in transparency practices. We used logistic regression adjusted by year of publication to test the relationship between transparency indicators and journal impact factor or received citations. We also used a random intercept generalized linear model to investigate the trends of transparency practices among different medical disciplines.

## Results

### General characteristics

As of March 16, 2024, EPMC contained 3,397,155 articles from the SCIE-indexed medical journals in total. Of these, 2,002,955 (59.0%) records had full text available for download. Of the articles, 40,307 (2.0%) were published before 2000, 812,146 (40.5%) in the 2010s, and 1,065,770 (53.2%) after 2020. The articles came from 3,458 journals, led by the International Journal of Environmental Research and Public Health (n=59,448, 3.0%), Frontiers in Immunology (n=32,224, 1.6%), and Medicine (Baltimore) (n=28,605, 1.4%). Among 450 publishers, Nature Publishing Group (465,741, 23.3%), Multidisciplinary Digital Publishing Institute (MDPI) (250,360, 12.5%), and Frontiers Media (167,114, 8.3%) had the highest number of papers. There were 261,803 (13.1%) reviews and 91,014 (4.5%) trials.

### Discipline-specific characteristics

On average, 89.2% (SD=8.60%) of the journals had at least one downloadable full-text in EPMC. Oncology (8.9%, n=219,797), Medicine, General & Internal (8.1%, n=199,331), and Medicine, Research & Experimental (7.2%, n=176,858) had the highest number of papers whereas Medicine, Legal (0.1%, n=1,231), Audiology & Speech-language Pathology (0.1%, n=1,571), and Medical Ethics (0.1%, n=2,959) had the lowest. For full-text availability in EPMC, Integrative & Complementary Medicine (88.8%, n=21,468), Tropical Medicine (84.6%, n=32,342), and Medicine, Research & Experimental (75.8%, n=176,858) led, while Substance Abuse (16.2%, n=3,623), Audiology & Speech-language Pathology (16.5%, n=1,571), and Peripheral Vascular Disease (24.8%, n=10,289) had the lowest availability (Appendix 2).

Medicine, Research & Experimental (13.7%, n=12,424), Medicine, General & Internal (12.5%, n=11,344), and Oncology (10.1%, n=9,214) had the highest number of trials. The highest number of reviews belonged to Oncology (12.1%, n=31,676), Medicine, General & Internal (11.1%, n=28,982), and Pharmacology & Pharmacy (10.1%, 26,493).

### Transparency practices overall

Of the analyzed full-text articles, 88.8% (95% CI: 88.8%-88.9%) disclosed a COI. Funding disclosures were present in 82.2% (82.1%-82.2%) of articles. Pre-publication registration was mentioned in 7.0% (6.9%-7.0%) and data sharing in 8.0% (8.0%-8.0%) of articles. Code sharing was reported in 1.5% (1.5%-1.5%) (Figure 1A - left). More than 78.3% mentioned at least two transparency indicators, while 0.02% complied with all five.

**Figure 1.**
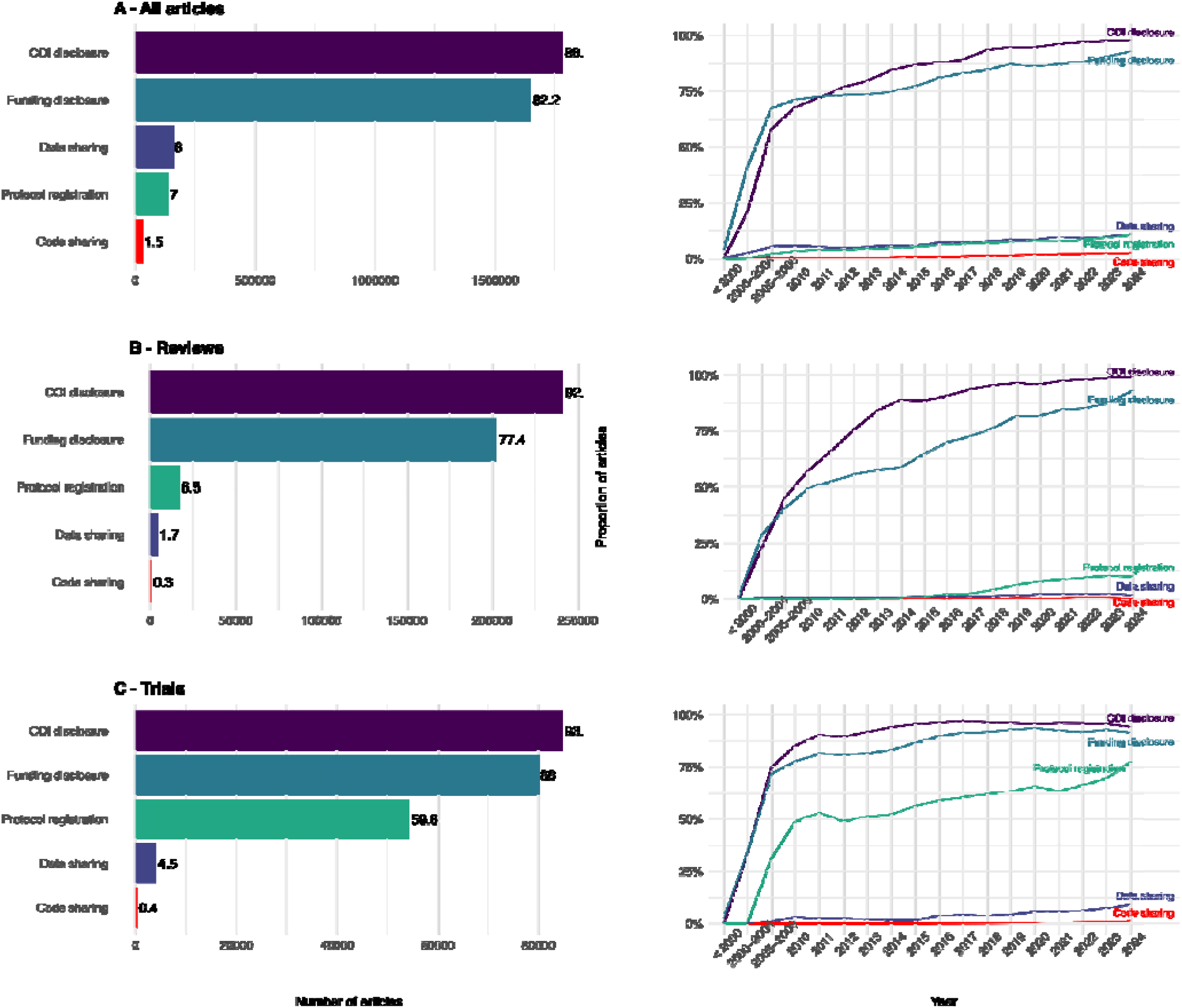
The proportion of 2,002,955 analyzed articles with transparency practices in total (A) and over time (B).

The left-hand side of Figures 1B and 1C show the adherence to transparency practices in reviews and trials. Reviews (92.2%, 92.1%-92.3%) and trials (93.1%, 92.9%-93.2%) had higher COI disclosure than all articles. Trials (88.0%, 87.8%-88.2%) mentioned funding disclosure more often than all articles but reviews less often (77.4%, 77.2%-77.5%). Protocol registration was mentioned in 59.6% (59.3%-60.0%) of the trials whereas reviews (6.5%, 6.5%-6.6%) mentioned preregistration less often compared to all articles. Data and code sharing were less often reported in reviews and trials than in all articles.

A visual analysis of the data since the end of the 2000s shows a steady increase in the proportion of articles reporting transparency practices (Figure 1, right). Pearson’s product-moment correlation between publication year and transparency practices was highest for funding disclosure (0.716) and lowest for code sharing (0.521). All the *P*-values were <0.001 (Appendix 1).

Articles published in journals with higher impact factors and articles receiving more citations had increased odds of funding disclosures, as well as data and code sharing (Figure 2A). The relationships of the journal impact factor and received citations with the odds of protocol registration were negative in all articles, and strongly negative in reviews. The relationships of the journal impact factor and received citations were more mixed.

**Figure 2.**
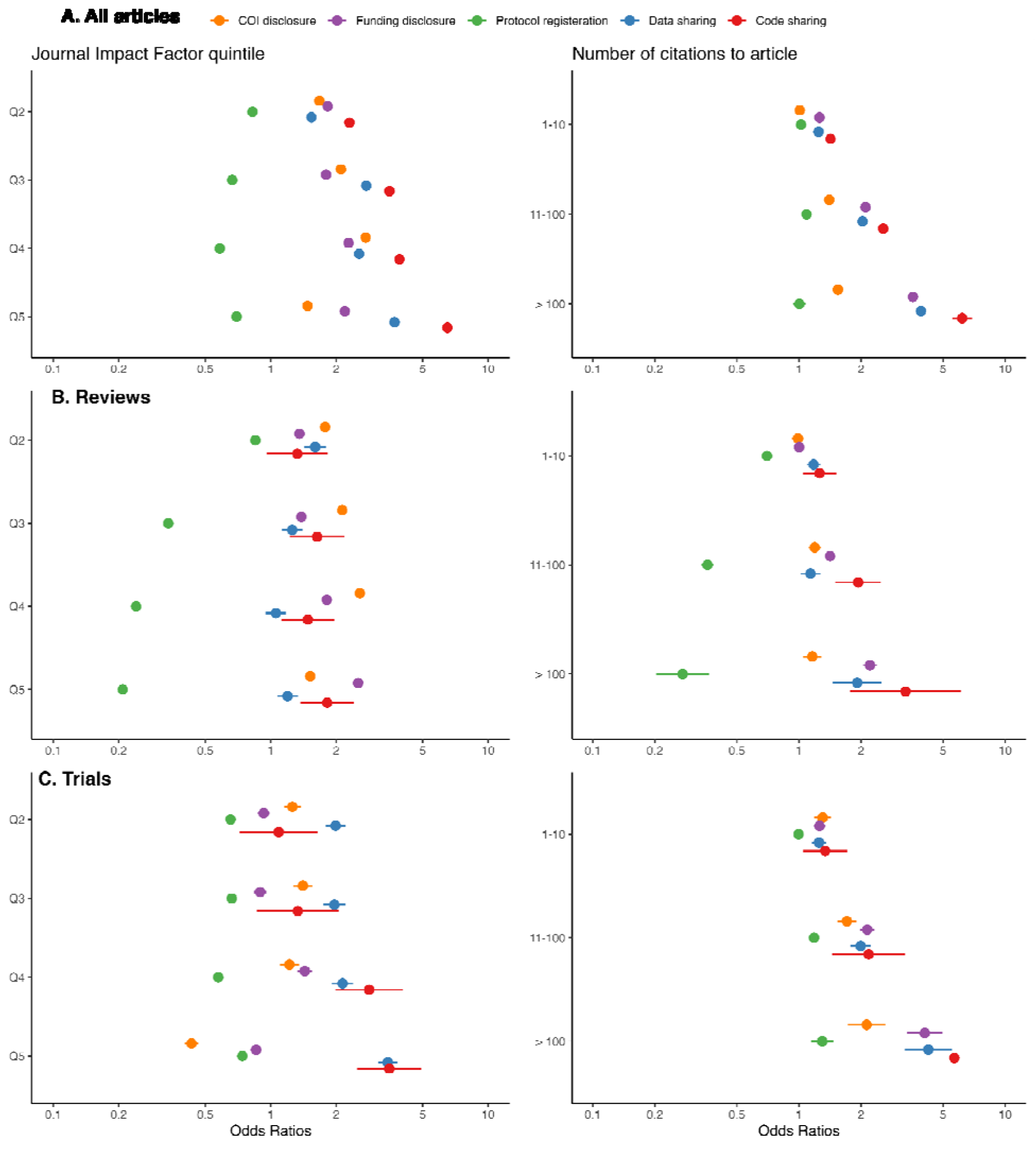
Transparency practices in research articles by Journal Impact Factor quintiles (A, lowest quintile, Q1 as the reference), and number of citations to article (B, zero citation as the reference). Odds ratios were adjusted for the year of article publication.

### Transparency practices by medical disciplines

#### Conflict of Interest Disclosure

The highest rates of COI disclosures were found in Rheumatology (98.1%), Primary Health Care (97.4%) and Emergency Medicine (96.8%). On the lower end, Medicine, Legal showed 61.6%, Toxicology 67.5%, and Neuroimaging 71.5% rate of COI disclosures (Appendix 3). The 2010s marked a period of increasing COI disclosures across most medical disciplines (Appendix 4).

#### Funding Disclosure

The highest rates of funding disclosure were found in Neuroimaging (94.9%), Materials Science, Biomaterials (94.0%), and Audiology & Speech-language Pathology (93.3%). In contrast, Medical Laboratory Technology reports 60.3%, Andrology 64.7%, and Critical Care Medicine 64.8% (Appendix 3). Funding disclosures, akin to COI disclosures, have shown an overall increase during the 2010s across various disciplines (Appendix 4).

#### Protocol Registration

Anesthesiology (34.5%) had the highest proportion of articles declaring protocol registration, followed by Rehabilitation (17.2%) and Critical Care Medicine (15.2%). Materials Science, Biomaterials (0.3%), Genetics & Heredity (0.9%), and Virology (1.2%) showed the lowest proportions (Appendix 3).

Anesthesiology, for example, has seen a steady increase, reaching up to 40%, contrasting with consistently low rates in disciplines like Immunology and Toxicology (Appendix 4).

#### Data Sharing

Data sharing was mentioned most frequently in Genetics & Heredity (37.9%), Neuroimaging (24.7%), and Virology (23.5%), while Surgery, Primary Health Care, and Orthopedics (all 1.6%) showed the lowest rates (Appendix 3). Although there has been an increase in data sharing in certain disciplines, such as Neuroimaging (approximately 80% in 2022), these trends are lower compared to COI and funding disclosures (Appendix 4).

#### Code Sharing

In code sharing, Neuroimaging (12.4%), Genetics & Heredity (7.6%), and Medical Informatics (6.3%) had the highest rates of code sharing, in contrast to negligible rates in Orthopedics, Nursing, and Integrative & Complementary Medicine (Appendix 3). Code sharing remains relatively low compared to the higher prevalence of COI and funding disclosures, though an increasing trend is observed in specific medical disciplines (Appendix 4).

The random intercept generalized linear mixed-effects logistic models examining transparency indicators over time among different disciplines showed the highest random effects variance for code sharing (1.347) and the lowest for funding disclosure (0.345). Full details are available in Appendix 1 and 5.

## Discussion

Our analyses of over two million full-text articles published in SCIE-indexed medical journals revealed high compliance with COI and funding disclosure in medical research since the early 2010s. However, data sharing, protocol registration, and code sharing remained relatively rare. A recent study highlights the critical role of best practices, such as pre-registration, in research (19). It demonstrates that adherence to these practices correlates with an 86% success rate in replication studies, significantly higher than the 50% success rate observed in some earlier replication efforts. Given the observed low rates of protocol registration, code sharing, and data sharing in our analysis, it is plausible that current research in these disciplines may face challenges in replicability, potentially falling short of the higher success rates associated with rigorous adherence to best practices.

To our knowledge, differences in transparency practices across a wide range of medical disciplines have not been studied before in this level of detail. Instead of focusing on a single transparency practice, such as data sharing, within one field of medicine or, e.g., its few highest-impact journals, the applied programmatic approach allowed us to estimate five transparency practices across a high number of studies across the range of medical disciplines (15). Previous research on data and code sharing has often taken a more generalized approach, focusing on overarching trends within the biomedical literature or narrowing their scope to specific disciplines or even individual journals (15). In contrast, our study delved into a detailed analysis, revealing substantial disparities, particularly in protocol registration and data or code sharing. These discrepancies likely mirror variations in research methodologies, reporting standards, publishing norms, peer review processes, and editorial practices prevalent across different medical domains. Conversely, our study indicates that COI and funding disclosures have been relatively prevalent since the mid-2000s. This suggests that achieving a high level of adherence to transparency practices could be attainable with concerted and universally applied efforts that have been behind promoting COI and funding disclosures (20). For instance, journals could adhere to Transparency and Openness Promotion Guidelines and guide authors accordingly (21). Recently, it was shown that it is possible to increase data and code sharing with such efforts, and thus subsequently increase the reproducibility of research substantially (22). Indeed, we believe that protocol registration, data and code sharing would become much more common if they were required for publication, similar to current requirements for COI and funding disclosures.

Even though we were able to use a large sample of articles and validated methods to estimate the prevalence of five transparency practices, our study also has some limitations. The analyzed articles represent, to some extent, a biased subset of all medical literature, even though earlier research has shown that there are no clear differences in these five transparency practices between articles with and without available full text in the EPMC (23). The applied methodology has not been validated for the whole time, the original validation covered the years 2015-2019 (13). We cannot be sure that it is equally valid/accurate within each of the 59 medical disciplines, because there may be some systematically different ways of registering protocols and sharing data or code, e.g. via some smaller and thus (uncoded) repositories/platforms. Furthermore, we did not manually evaluate the validity or accuracy of the disclosures, protocols, or data/code availability statements, but other studies have found similar results: these are frequently suboptimal (24,25). That is one reason why we are conducting further research to investigate findability, accessibility, interoperability, and reusability datasets from the articles reporting sharing data (26). In a similar study on COVID-19 research data availability, we found that among 8,015 articles detected by the *rtransparent* tool as having shared their data in a general repository, 5,700 (71.1%) actually had available data (27). It is also important to note that the articles in our sample included a wide range of research, some of which may not have necessitated using any data or code. Consequently, we acknowledge that achieving a 100% rate in data and code sharing would not be a realistic expectation. For instance, disciplines such as “Medical Ethics” or “Medicine, Legal” may predominantly comprise qualitative research papers, which may not inherently involve creating or utilizing datasets or codes. Instead, in disciplines like “Genetics & Heredity” data-intensive research is likely more common. Additionally, the issue of protocol registration introduces some complexity because there is no consensus regarding the necessity of pre-registration for certain study types, such as explorative (as opposed to hypothesis-testing) or qualitative research. These discrepancies likely drive the observed variations across the 59 distinct medical disciplines. Consequently, it is essential to interpret the results with caution and make comparisons with an understanding of these inherent variations across the diverse spectrum of medical research.

Compared to all articles with available full-text in the EPMC database (13), the analyzed subsample of articles from SCIE-indexed medical journals showed higher prevalence of COI and funding disclosures and protocol registration, but equal prevalence data and code-sharing. It seems these disparities primarily stem from historical trends. Specifically, the SCIE-indexed articles exhibited higher levels of transparency practices in earlier years. Interestingly, in 2020, the sample comprising all articles with available full-text in the EPMC database demonstrated greater data and code sharing, but lower adherence to protocol registration, COI disclosures, and funding disclosures when compared to our findings of the SCIE-indexed articles in 2024 (in parenthesis): 15% (11%) for data sharing, 3% (2.5%) for code sharing, 90% (97%) for COI disclosures, 85% (92%) for funding disclosures, and 5% (10%) for protocol registration. Menke et al. have also investigated the presence of protocols and data and code sharing in all articles with full-text in the EPMC database with another automated tool (28). They found a similar constant trend in code sharing that we did. However, they found a more pronounced increase in and higher overall adherence to protocol registration (19% vs. 9%) and data sharing (17% vs. 9%) in 2020 than our findings indicate, potentially attributable to differences in these automated methods.

In conclusion, our study reveals that key transparency practices such as data sharing, protocol registration, and especially code sharing continue to be notably scarce in medical research. While adherence to COI and funding disclosures is commendable, the limited adoption of these other crucial transparency practices across diverse medical disciplines remains a significant concern. The recent findings highlighting a high replication success rate with rigorous transparency, underscore the vital need for the universal adoption of such practices (19,22). We urge researchers, journal editors, and policymakers to advance these practices by advocating for the standardization of protocol registration and open data/code sharing across all medical disciplines. Such a collective commitment is essential to enhance the integrity, reliability, and impact of medical research, ultimately benefiting the global health community.

## Data Availability

All the code and data associated with the study were shared through both its OSF repository (https://osf.io/zbc6p/) and GitHub (https://github.com/choxos/medical-transparency) when the manuscript was submitted.

## Acknowledgments

The computational analyses were performed on servers provided by UEF Bioinformatics Center, University of Eastern Finland, Finland. Uribe was supported by the European Union’s Horizon 2020 grant 857287 for the Baltic Biomaterials Centre of Excellence, Headquarters at Riga Technical University, Riga, Latvia.

## Notes

**Conflict of interest disclosure**: The authors have no conflicts of interest to disclose.

### Competing Interest Statement

The authors have declared no competing interest.

### Funding Statement

This study did not receive any funding.

### Summary of Updates

We changed the inclusion criteria to only research articles and reviews. Also, we added analyses based on article type (all articles, reviews, and trials).

