## Appendix 1 for "Research Transparency in 59 Disciplines of Clinical Medicine: A Meta-Research Study"

Ahmad Sofi-Mahmudi

#### Table of contents


**1 Aim** 2

**2 Results** 2

2.1 **General characteristics** 3
2.2 **Category-specific characteristics** 9
2.2.1 Appendix 2 10
2.3 **Overall adherence to transparency practices** 13
2.3.1 All articles 13
2.4 Adherence by number of practices 15
2.4.1 Reviews 16
2.4.2 Trials 18
2.5 Transparency practices by fields over time 20
2.5.1 Overall trend 20
2.5.2 Appendix 4: Yearly trends for each field 33
2.5.3 Appendix 3 39
2.6 Association with impact factor and number of citations 42
2.6.1 Figure 2 42
2.7 Publisher differences 45
2.7.1 Conflict of interest disclosure 45
2.7.2 Funding disclosure 47
2.7.3 Protocol registration 48
2.7.4 Data sharing 49
2.7.5 Code sharing 49

### 1 Aim

We aimed to assess the adherence to five transparent practices (data availability, code availability, protocol registration and conflicts of interest (COI), and funding disclosures) from open-access articles published in medical journals.

### 2 Results

First, loading the needed packages:

```
pacman::p_load(dplyr,
               ggplot2,
               knitr,
               gtsummary,
               tidyr,
               lubridate,
               forcats,
               DiagrammeR,
               tibble,
               ggrepel,
               ggpubr,
               epiR,
               here,
               lme4,
               nortest,
               stringr,
               expss,
               dvmisc,
               sjPlot,
               eply)
```

And then, loading the datasets:

```
transparency = read.csv("data/medicaltransparency_opendata.csv")
transparency = transparency %>% filter(type %in% c("research-article", "review-article", "sys
```

Now, let's create a dataset without the duplicates:

```
unique_transparency = transparency[!duplicated(transparency$pmid), ]
```

#### 2.1 General characteristics

Number of all papers (open access and non open access), open-access papers and open-access percentage.

First, we delete duplicated ISSNs to capture the true number of all articles (without any duplication):

```
# ISSNs = read.csv("data/journals.csv")
# ISSNs = ISSNs %>% filter(Category != "MULTIDISCIPLINARY SCIENCES - SCIE")
# ISSNs_unique = ISSNs[!duplicated(ISSNs$ISSN), ]
# ISSNs_unique = ISSNs_unique %>% mutate(search.term = paste0("ISSN:", ISSN))
# ISSNsQuery_unique = ISSNs_unique %>%
#        group_by(Category) %>% summarize(query = paste(search.term,
#                                                      collapse = " OR "))
# ISSNsQuery_unique = as.data.frame(ISSNsQuery_unique)

# hits_list_unique = data.frame()
# hits_list_unique = sapply(ISSNsQuery_unique$query, function(issn) {
#        search_string = paste0(
#                "(",
#                issn,
#                ") ",
#                'AND (SRC:"MED")
#    AND (LANG:"eng" OR LANG:"en" OR LANG:"us")
#    AND (FIRST_PDATE:[1990-01-01 TO 2024-03-16])
#    AND (PUB_TYPE:"research-article" OR PUB_TYPE:"review-article" OR PUB_TYPE:"systematic-r
#        )
#        epmc_hits(query = search_string)
#})

# hits_list_unique = as.data.frame(hits_list_unique)
# rownames(hits_list_unique) = ISSNsQuery$Category
# write.csv(hits_list_unique, "data/hits_list_unique.csv")
hits_list_unique = read.csv("data/hits_list_unique.csv")
```

Now, we do the same but with duplicates to have the exact number of articles for each category:

```
# ISSNs = ISSNs %>% mutate(search.term = paste0("ISSN:", ISSN))
# ISSNsQuery = ISSNs %>%
#        group_by(Category) %>% summarize(query = paste(search.term,
```

```
#                                                          collapse = " OR "))
# ISSNsQuery = as.data.frame(ISSNsQuery)

# hits_list = data.frame()
# hits_list = sapply(ISSNsQuery$query, function(issn) {
#        search_string = paste0(
#                "(",
#                issn,
#                ") ",
#                'AND (SRC:"MED")
#    AND (LANG:"eng" OR LANG:"en" OR LANG:"us")
#    AND (FIRST_PDATE:[1990-01-01 TO 2024-03-16])
#    AND (PUB_TYPE:"research-article" OR PUB_TYPE:"review-article" OR PUB_TYPE:"systematic-re
#        )
#        epmc_hits(query = search_string)
#})

# hits_list = as.data.frame(hits_list)
# rownames(hits_list) = ISSNsQuery$Category
# write.csv(hits_list, "data/hits_list.csv")

hits_list = read.csv("data/hits_list.csv")
```

Now, calculating open-access percentage:

```
kable(data.frame(hits_all = sum(hits_list_unique$hits_list_unique),
           hits_oa = nrow(unique_transparency),
           oa_percentage = round((nrow(unique_transparency)/sum(hits_list_unique$hits_list_un
```

| hits_all | hits_oa | oa_percentage |
|---|---|---|
| 3397155 | 2002955 | 59 |

Let's take a look at the number of papers published in each year and overall. To do so, first, we add the real publication year and month to the datasets. The real publication year/month is the year/month the paper was first appear online stored in firstPublicationDate column.

```
unique_transparency = unique_transparency %>%
  mutate(year_firstpub = year(
    as.POSIXlt(firstPublicationDate,
               format = "%Y-%m-%d")),
```

```r
    month_firstpub = month(
      as.POSIXlt(firstPublicationDate,
               format = "%Y-%m-%d")
    )
    )

unique_transparency = unique_transparency %>%
        mutate(pubYear_modified =
                       ifelse(year_firstpub < 2000, "< 2000",
                     ifelse((year_firstpub >= 2000) & (year_firstpub < 2005), "2000-2004",
                     ifelse((year_firstpub >= 2005) & (year_firstpub < 2010), "2005-2009",
                     year_firstpub))))

transparency = transparency %>%
  mutate(year_firstpub = year(
    as.POSIXlt(firstPublicationDate,
               format = "%Y-%m-%d")),
    month_firstpub = month(
      as.POSIXlt(firstPublicationDate,
               format = "%Y-%m-%d")
    )
    )

transparency = transparency %>%
        mutate(pubYear_modified =
                       ifelse(year_firstpub < 2000, "< 2000",
                     ifelse((year_firstpub >= 2000) & (year_firstpub < 2005), "2000-2004",
                     ifelse((year_firstpub >= 2005) & (year_firstpub < 2010), "2005-2009",
                     year_firstpub))))
```

Now, the number of open access papers per year:

```r
unique_transparency %>% tbl_summary(include = c(pubYear_modified),
                         digits = list(all_categorical() ~ c(0, 1)))
```

```
Table printed with `knitr::kable()`, not {gt}. Learn why at
https://www.danieldsjoberg.com/gtsummary/articles/rmarkdown.html
To suppress this message, include `message = FALSE` in code chunk header.
```

| **Characteristic** | **N = 2,002,955** |
| --- | --- |
| pubYear_modified | |
| < 2000 | 40,307 (2.0%) |
| 2000–2004 | 16,711 (0.8%) |
| 2005–2009 | 68,021 (3.4%) |
| 2010 | 30,773 (1.5%) |
| 2011 | 37,739 (1.9%) |
| 2012 | 47,077 (2.4%) |
| 2013 | 62,166 (3.1%) |
| 2014 | 75,961 (3.8%) |
| 2015 | 84,894 (4.2%) |
| 2016 | 91,473 (4.6%) |
| 2017 | 105,762 (5.3%) |
| 2018 | 126,410 (6.3%) |
| 2019 | 149,891 (7.5%) |
| 2020 | 213,102 (10.6%) |
| 2021 | 276,896 (13.8%) |
| 2022 | 303,395 (15.1%) |
| 2023 | 238,246 (11.9%) |
| 2024 | 34,131 (1.7%) |

Paper types frequency:

```
unique_transparency %>% tbl_summary(include = c(type),
                                    digits = list(all_categorical() ~ c(0, 1)))
```

```
Table printed with `knitr::kable()`, not {gt}. Learn why at
https://www.danieldsjoberg.com/gtsummary/articles/rmarkdown.html
To suppress this message, include `message = FALSE` in code chunk header.
```

| **Characteristic** | **N = 2,002,955** |
| --- | --- |
| type | |
| research-article | 1,741,152 (86.9%) |
| review-article | 258,470 (12.9%) |
| systematic-review | 3,333 (0.2%) |

Now, comparing journals:

```
set.seed(1280)

unique_transparency %>%
        select(journalTitle,
               is_coi_pred,
               is_fund_pred,
               is_register_pred,
               is_open_data,
               is_open_code) %>%
        mutate(journalTitle = fct_lump(journalTitle, n = 5)) %>%
        tbl_summary(by = journalTitle,
                    percent = "column",
                    label = c(is_coi_pred ~ "COI disclosure",
                              is_fund_pred ~ "Funding disclosure",
                              is_register_pred ~ "Protocol registration",
                              is_open_data ~ "Data sharing",
                              is_open_code ~ "Code sharing"),
                    digits = list(all_categorical() ~ c(0, 1))) %>%
        add_p(test.args = all_tests("fisher.test") ~ list(simulate.p.value=TRUE)) %>%
        as_flex_table()
```

| **Characteristic** | **Front Immunol**, N = 32,224[1] | **Int J Environ Res Public Health**, N = 59,448[1] |
|---|---|---|
| COI disclosure | 32,174 (99.8%) | 58,530 (98.5%) |
| Funding disclosure | 29,358 (91.1%) | 57,781 (97.2%) |
| Protocol registration | 606 (1.9%) | 2,581 (4.3%) |
| Data sharing | 3,204 (9.9%) | 1,881 (3.2%) |
| Code sharing | 400 (1.2%) | 320 (0.5%) |

[1]n (%)

[2]Pearson's Chi-squared test

Publishers:

```
set.seed(1280)

unique_transparency %>%
        select(scimago_publisher,
               is_coi_pred,
```

```
            is_fund_pred,
            is_register_pred,
            is_open_data,
            is_open_code) %>%
    mutate(scimago_publisher = fct_lump(scimago_publisher, n = 5)) %>%
    tbl_summary(by = scimago_publisher,
                percent = "column",
                label = c(is_coi_pred ~ "COI disclosure",
                          is_fund_pred ~ "Funding disclosure",
                          is_register_pred ~ "Protocol registration",
                          is_open_data ~ "Data sharing",
                          is_open_code ~ "Code sharing"),
                digits = list(all_categorical() ~ c(0, 1))) %>%
    add_p(test.args = all_tests("fisher.test") ~ list(simulate.p.value=TRUE)) %>%
    as_flex_table()
```

```
190773 observations missing `scimago_publisher` have been removed. To include these observati
```

| **Characteristic** | **Elsevier**, N = 86,540[1] | **Frontiers Media S.A.**, N = 167,114[1] | **John Wiley &** |
|---|---|---|---|
| COI disclosure | 76,044 (87.9%) | 166,674 (99.7%) | 101 |
| Funding disclosure | 73,953 (85.5%) | 142,884 (85.5%) | 95 |
| Protocol registration | 9,143 (10.6%) | 5,194 (3.1%) | 8 |
| Data sharing | 6,326 (7.3%) | 11,831 (7.1%) | 11 |
| Code sharing | 1,374 (1.6%) | 3,474 (2.1%) | 1 |

[1]n (%)

[2]Pearson's Chi-squared test

What about article types?

```
set.seed(1280)

unique_transparency %>%
    select(type,
           is_coi_pred,
           is_fund_pred,
```

```
            is_register_pred,
            is_open_data,
            is_open_code) %>%
    mutate(type = fct_lump(type, n = 4)) %>%
    tbl_summary(by = type,
                percent = "column",
                label = c(is_coi_pred ~ "COI disclosure",
                          is_fund_pred ~ "Funding disclosure",
                          is_register_pred ~ "Protocol registration",
                          is_open_data ~ "Data sharing",
                          is_open_code ~ "Code sharing"),
                digits = list(all_categorical() ~ c(0, 1))) %>%
    add_p(test.args = all_tests("fisher.test") ~ list(simulate.p.value=TRUE)) %>%
    as_flex_table()
```

| **Characteristic** | **research-article**, N = 1,741,152[1] | **review-article**, N = 258,470[1] | **systematic** |
|---|---|---|---|
| COI disclosure | 1,538,157 (88.3%) | 237,981 (92.1%) | [illegible] |
| Funding disclosure | 1,443,743 (82.9%) | 199,472 (77.2%) | [illegible] |
| Protocol registration | 122,617 (7.0%) | 15,183 (5.9%) | [illegible] |
| Data sharing | 156,064 (9.0%) | 4,014 (1.6%) | [illegible] |
| Code sharing | 29,068 (1.7%) | 707 (0.3%) | |

[1]n (%)

[2]Pearson's Chi-squared test

#### 2.2 Category-specific characteristics

Top categories in terms of number of papers

Top 3 highest:

```
transparency %>% group_by(category) %>% summarise(n = n(), p = round(n/nrow(transparency)*10
```

```
# A tibble: 3 x 3
  category                              n     p
  <chr>                             <int> <dbl>
1 Oncology                         219797  8.9
2 Medicine, General & Internal     199331  8.07
3 Medicine, Research & Experimental 176858  7.16
```

Top 3 lowest:

```
transparency %>% group_by(category) %>% summarise(n = n(), p = round(n/nrow(transparency)*10
```

```
# A tibble: 3 x 3
  category                                  n     p
  <chr>                                 <int> <dbl>
1 Medicine, Legal                        1231  0.05
2 Audiology & Speech-language Pathology  1571  0.06
3 Medical Ethics                         2959  0.12
```

Fields with the highest number of trials and reviews

Trials:

```
transparency %>% filter(is_trial == T) %>% group_by(category) %>% summarise(n = n(), p=round
```

```
# A tibble: 3 x 3
  category                             n     p
  <chr>                            <int> <dbl>
1 Medicine, Research & Experimental 12424  13.6
2 Medicine, General & Internal      11344  12.5
3 Oncology                           9214  10.1
```

Reviews:

```
transparency %>% filter(is_review == T) %>% group_by(category) %>% summarise(n = n(), p=roun
```

```
# A tibble: 3 x 3
  category                        n     p
  <chr>                       <int> <dbl>
1 Oncology                    31676  12.1
2 Medicine, General & Internal 28982  11.1
3 Pharmacology & Pharmacy     26493  10.1
```

##### 2.2.1 Appendix 2

Column 1: Number of journals in each category

```
ISSNs = read.csv("data/journals.csv")
ISSNs = ISSNs %>% filter(Category != "MULTIDISCIPLINARY SCIENCES - SCIE")
appendix2 = data.frame()
appendix2 = ISSNs %>% group_by(Category) %>% summarise(all_journals = n())
```

Column 2: Available journals in each category based on ISSN

```
appendix2$available_journals_issn = (transparency %>% group_by(category) %>% summarise(availa
```

Column 3: Percentage based on column 2

```
appendix2$availability_percentage_issn = appendix2$available_journals_issn/appendix2$all_jour
```

Column 4: Available journals in each category based on journal title

```
appendix2$available_journals_name = (transparency %>% group_by(category) %>% summarise(availa
```

Column 5: Percentage based on column 4

```
appendix2$availability_percentage_name = round(appendix2$available_journals_name/appendix2$al
```

Column 6: All articles

```
appendix2$all_articles_20240316 = hits_list$hits_list
```

Column 7: Open-access articles

```
appendix2$oa_articles_20240316 = table(transparency$category)
```

Column 8: Open-access percentage

```
appendix2$percentage_oa = round(appendix2$oa_articles_20240316/appendix2$all_articles_202403
```

Now, we can save it:

```
# write.csv(appendix2, "appendix/Appendix2.csv", row.names = F)
```

Open-access availability based on name:

```
c(mean(appendix2$availability_percentage_name), sd(appendix2$availability_percentage_name))
```

```
[1] 89.178305  8.596136
```

Top 3 categories with the most open-access availability:

```
appendix2 %>% select(Category, oa_articles_20240316, percentage_oa) %>% arrange(desc(percenta
```

```
# A tibble: 3 x 3
  Category                                      oa_articles_20240316 percentage_oa
  <chr>                                         <table[1d]>          <table[1d]>
1 INTEGRATIVE & COMPLEMENTARY MEDICINE - SCIE    21468               88.81
2 TROPICAL MEDICINE - SCIE                       32342               84.58
3 MEDICINE, RESEARCH & EXPERIMENTAL - SCIE      176858               75.84
```

Top 3 categories with the least open-access availability:

```
appendix2 %>% select(Category, oa_articles_20240316, percentage_oa) %>% arrange(percentage_oa
```

```
# A tibble: 3 x 3
  Category                                      oa_articles_20240316 percentage_oa
  <chr>                                         <table[1d]>          <table[1d]>
1 SUBSTANCE ABUSE - SCIE                         3623                16.22
2 AUDIOLOGY & SPEECH-LANGUAGE PATHOLOGY - SC~    1571                16.48
3 PERIPHERAL VASCULAR DISEASE - SCIE            10289                24.76
```

And categories:

```
set.seed(1280)

transparency %>%
        select(category,
               is_coi_pred,
               is_fund_pred,
               is_register_pred,
               is_open_data,
               is_open_code) %>%
        mutate(category = fct_lump(category, n = 5)) %>%
        tbl_summary(by = category,
```

```
                    percent = "column",
                    label = c(is_coi_pred ~ "COI disclosure",
                              is_fund_pred ~ "Funding disclosure",
                              is_register_pred ~ "Protocol registration",
                              is_open_data ~ "Data sharing",
                              is_open_code ~ "Code sharing"),
                    digits = list(all_categorical() ~ c(0, 1))) %>%
        add_p(test.args = all_tests("fisher.test") ~ list(simulate.p.value=TRUE)) %>%
        as_flex_table()
```

| Characteristic | **Medicine, General & Internal**, N = 199,331[1] | **Medicine, Research & Experin** |
|---|---|---|
| COI disclosure | 180,703 (90.7%) | 138,666 (78.4% |
| Funding disclosure | 159,090 (79.8%) | 140,803 (79.6% |
| Protocol registration | 25,679 (12.9%) | 12,689 (7.2%) |
| Data sharing | 8,795 (4.4%) | 13,920 (7.9%) |
| Code sharing | 1,416 (0.7%) | 1,516 (0.9%) |

[1]n (%)

[2]Pearson's Chi-squared test

#### 2.3 Overall adherence to transparency practices

##### 2.3.1 All articles

Number and percentage:

```
kable(rbind(
        COI = data.frame(number = length(unique_transparency$is_coi_pred[unique_transparency$
           percentage = round(length(unique_transparency$is_coi_pred[unique_transparency$is_c
           ),
         Fund = data.frame(number = length(unique_transparency$is_fund_pred[unique_transparen
           percentage = round(length(unique_transparency$is_fund_pred[unique_transparency$is_
           ),
        Register = data.frame(number = length(unique_transparency$is_register_pred[unique_tra
           percentage = round(length(unique_transparency$is_register_pred[unique_transparency
           ),
        Data = data.frame(number = length(unique_transparency$is_open_data[unique_transparenc
           percentage = round(length(unique_transparency$is_open_data[unique_transparency$is_
```

```
        ),
      Code = data.frame(number = length(unique_transparency$is_open_code[unique_transparenc
        percentage = round(length(unique_transparency$is_open_code[unique_transparency$is_
        )
        ))
```

| | number | percentage |
|---|---|---|
| COI | 1779458 | 88.8 |
| Fund | 1646303 | 82.2 |
| Register | 139764 | 7.0 |
| Data | 160397 | 8.0 |
| Code | 29786 | 1.5 |

And CIs:

```
kable(rbind(COI=round(epi.prev(pos = length(unique_transparency$is_coi_pred[unique_transparen
        tested = nrow(unique_transparency),
        se = 0.992,
        sp = 0.995)$ap,
      1),
      Funding=round(epi.prev(pos = length(unique_transparency$is_fund_pred[unique_transparenc
        tested = nrow(unique_transparency),
        se = 0.997,
        sp = 0.981)$ap,
      1),
      Protocol=round(epi.prev(pos = length(unique_transparency$is_register_pred[unique_transp
        tested = nrow(unique_transparency),
        se = 0.955,
        sp = 0.997)$ap,
      1),
      Data=round(epi.prev(pos = length(unique_transparency$is_open_data[unique_transparency$i
        tested = nrow(unique_transparency),
        se = 0.758,
        sp = 0.986)$ap,
      1),
      Code=round(epi.prev(pos = length(unique_transparency$is_open_code[unique_transparency$i
        tested = nrow(unique_transparency),
        se = 0.587,
        sp = 0.997)$ap,
      1)))
```

| | est | lower | upper |
|---|---|---|---|
| COI | 88.8 | 88.8 | 88.9 |
| Funding | 82.2 | 82.1 | 82.2 |
| Protocol | 7.0 | 6.9 | 7.0 |
| Data | 8.0 | 8.0 | 8.0 |
| Code | 1.5 | 1.5 | 1.5 |

#### 2.4 Adherence by number of practices

```
unique_transparency = unique_transparency %>% mutate(sumOfIndicators = rowSums(unique_transpa
```

Number of papers with each number of TRUE indicators:

```
c(five_ind = nrow(filter(unique_transparency, sumOfIndicators == 5)),
  four_ind = nrow(filter(unique_transparency, sumOfIndicators == 4)),
  three_ind = nrow(filter(unique_transparency, sumOfIndicators == 3)),
  two_ind = nrow(filter(unique_transparency, sumOfIndicators == 2)),
  one_ind = nrow(filter(unique_transparency, sumOfIndicators == 1)),
  zero_ind = nrow(filter(unique_transparency, sumOfIndicators == 0)))
```

```
 five_ind  four_ind three_ind   two_ind   one_ind  zero_ind
      419     22512    247187   1298604    324796    109437
```

Percentage of papers with each number of TRUE indicators:

```
c(five_ind = round(nrow(filter(unique_transparency, sumOfIndicators == 5))/nrow(unique_transp
  four_ind = round(nrow(filter(unique_transparency, sumOfIndicators == 4))/nrow(unique_transp
  three_ind = round(nrow(filter(unique_transparency, sumOfIndicators == 3))/nrow(unique_trans
  two_ind = round(nrow(filter(unique_transparency, sumOfIndicators == 2))/nrow(unique_transpa
  one_ind = round(nrow(filter(unique_transparency, sumOfIndicators == 1))/nrow(unique_transpa
  zero_ind = round(nrow(filter(unique_transparency, sumOfIndicators == 0))/nrow(unique_transp
```

```
 five_ind  four_ind three_ind   two_ind   one_ind  zero_ind
   0.0209    1.1000   12.3000   64.8000   16.2000    5.5000
```

##### 2.4.1 Reviews

Number and percentage:

```
reviews = unique_transparency %>% filter(is_review == T)

kable(rbind(
        COI = data.frame(number = length(reviews$is_coi_pred[reviews$is_coi_pred == TRUE]),
           percentage = round(length(reviews$is_coi_pred[reviews$is_coi_pred == TRUE])/nrow(r
           ),
         Fund = data.frame(number = length(reviews$is_fund_pred[reviews$is_fund_pred == TRUE]
           percentage = round(length(reviews$is_fund_pred[reviews$is_fund_pred == TRUE])/nrow
           ),
        Register = data.frame(number = length(reviews$is_register_pred[reviews$is_register_pr
           percentage = round(length(reviews$is_register_pred[reviews$is_register_pred == TRU
           ),
        Data = data.frame(number = length(reviews$is_open_data[reviews$is_open_data == TRUE])
           percentage = round(length(reviews$is_open_data[reviews$is_open_data == TRUE])/nrow
           ),
        Code = data.frame(number = length(reviews$is_open_code[reviews$is_open_code == TRUE])
           percentage = round(length(reviews$is_open_code[reviews$is_open_code == TRUE])/nrow
           )
           ))
```

| | number | percentage |
|---|---|---|
| COI | 241301 | 92.2 |
| Fund | 202560 | 77.4 |
| Register | 17147 | 6.5 |
| Data | 4333 | 1.7 |
| Code | 718 | 0.3 |

And CIs:

```
kable(rbind(COI=round(epi.prev(pos = length(reviews$is_coi_pred[reviews$is_coi_pred == TRUE])
         tested = nrow(reviews),
         se = 0.992,
         sp = 0.995)$ap,
      1),
      Funding=round(epi.prev(pos = length(reviews$is_fund_pred[reviews$is_fund_pred == TRUE])
         tested = nrow(reviews),
         se = 0.997,
```

```
          sp = 0.981)$ap,
       1),
       Protocol=round(epi.prev(pos = length(reviews$is_register_pred[reviews$is_register_pred
          tested = nrow(reviews),
          se = 0.955,
          sp = 0.997)$ap,
       1),
       Data=round(epi.prev(pos = length(reviews$is_open_data[reviews$is_open_data == TRUE]),
          tested = nrow(reviews),
          se = 0.758,
          sp = 0.986)$ap,
       1),
       Code=round(epi.prev(pos = length(reviews$is_open_code[reviews$is_open_code == TRUE]),
          tested = nrow(reviews),
          se = 0.587,
          sp = 0.997)$ap,
       1)))
```

```
Warning in epi.prev(pos = length(reviews$is_open_code[reviews$is_open_code == :
Apparent prevalence is less than (1 - Sp). Rogan Gladen estimate of true
prevalence invalid.
```

| | est | lower | upper |
|---|---|---|---|
| COI | 92.2 | 92.1 | 92.3 |
| Funding | 77.4 | 77.2 | 77.5 |
| Protocol | 6.5 | 6.5 | 6.6 |
| Data | 1.7 | 1.6 | 1.7 |
| Code | 0.3 | 0.3 | 0.3 |

Percentage of papers with each number of TRUE indicators:

```
c(five_ind = round(nrow(filter(reviews, sumOfIndicators == 5))/nrow(reviews)*100, 4),
  four_ind = round(nrow(filter(reviews, sumOfIndicators == 4))/nrow(reviews)*100, 1),
  three_ind = round(nrow(filter(reviews, sumOfIndicators == 3))/nrow(reviews)*100, 1),
  two_ind = round(nrow(filter(reviews, sumOfIndicators == 2))/nrow(reviews)*100, 1),
  one_ind = round(nrow(filter(reviews, sumOfIndicators == 1))/nrow(reviews)*100, 1),
  zero_ind = round(nrow(filter(reviews, sumOfIndicators == 0))/nrow(reviews)*100, 1))
```

```
 five_ind  four_ind three_ind   two_ind   one_ind  zero_ind
   0.0118    0.4000    6.7000   67.8000   20.5000    4.5000
```

##### 2.4.2 Trials

Number and percentage:

```
trials = unique_transparency %>% filter(is_trial == T)

kable(rbind(
        COI = data.frame(number = length(trials$is_coi_pred[trials$is_coi_pred == TRUE]),
           percentage = round(length(trials$is_coi_pred[trials$is_coi_pred == TRUE])/nrow(tr
           ),
         Fund = data.frame(number = length(trials$is_fund_pred[trials$is_fund_pred == TRUE])
           percentage = round(length(trials$is_fund_pred[trials$is_fund_pred == TRUE])/nrow(
           ),
        Register = data.frame(number = length(trials$is_register_pred[trials$is_register_pre
           percentage = round(length(trials$is_register_pred[trials$is_register_pred == TRUE
           ),
        Data = data.frame(number = length(trials$is_open_data[trials$is_open_data == TRUE]),
           percentage = round(length(trials$is_open_data[trials$is_open_data == TRUE])/nrow(
           ),
        Code = data.frame(number = length(trials$is_open_code[trials$is_open_code == TRUE]),
           percentage = round(length(trials$is_open_code[trials$is_open_code == TRUE])/nrow(
           )
           ))
```

| | number | percentage |
|---|---|---|
| COI | 84689 | 93.1 |
| Fund | 80102 | 88.0 |
| Register | 54282 | 59.6 |
| Data | 4118 | 4.5 |
| Code | 335 | 0.4 |

And CIs:

```
kable(rbind(COI=round(epi.prev(pos = length(trials$is_coi_pred[trials$is_coi_pred == TRUE]),
         tested = nrow(trials),
         se = 0.992,
         sp = 0.995)$ap,
      1),
      Funding=round(epi.prev(pos = length(trials$is_fund_pred[trials$is_fund_pred == TRUE]),
         tested = nrow(trials),
         se = 0.997,
```

```
          sp = 0.981)$ap,
      1),
      Protocol=round(epi.prev(pos = length(trials$is_register_pred[trials$is_register_pred ==
          tested = nrow(trials),
          se = 0.955,
          sp = 0.997)$ap,
      1),
      Data=round(epi.prev(pos = length(trials$is_open_data[trials$is_open_data == TRUE]),
          tested = nrow(trials),
          se = 0.758,
          sp = 0.986)$ap,
      1),
      Code=round(epi.prev(pos = length(trials$is_open_code[trials$is_open_code == TRUE]),
          tested = nrow(trials),
          se = 0.587,
          sp = 0.997)$ap,
      1)))
```

| | est | lower | upper |
|---|---|---|---|
| COI | 93.1 | 92.9 | 93.2 |
| Funding | 88.0 | 87.8 | 88.2 |
| Protocol | 59.6 | 59.3 | 60.0 |
| Data | 4.5 | 4.4 | 4.7 |
| Code | 0.4 | 0.3 | 0.4 |

Percentage of papers with each number of TRUE indicators:

```
c(five_ind = round(nrow(filter(trials, sumOfIndicators == 5))/nrow(trials)*100, 4),
  four_ind = round(nrow(filter(trials, sumOfIndicators == 4))/nrow(trials)*100, 1),
  three_ind = round(nrow(filter(trials, sumOfIndicators == 3))/nrow(trials)*100, 1),
  two_ind = round(nrow(filter(trials, sumOfIndicators == 2))/nrow(trials)*100, 1),
  one_ind = round(nrow(filter(trials, sumOfIndicators == 1))/nrow(trials)*100, 1),
  zero_ind = round(nrow(filter(trials, sumOfIndicators == 0))/nrow(trials)*100, 1))
```

```
 five_ind  four_ind three_ind   two_ind   one_ind  zero_ind
   0.0934    3.0000   53.3000   32.4000    8.6000    2.7000
```

##### 2.5 Transparency practices by fields over time

###### 2.5.1 Overall trend

First, we calculate the proportion of adherence to each domain overall and yearly adherence to each domain:

```
# Overall
proportions = unique_transparency %>%
        summarise("COI disclosure" = sum(is_coi_pred == TRUE),
                  "Funding disclosure" = sum(is_fund_pred == TRUE),
                  "Protocol registration" = sum(is_register_pred == TRUE),
                  "Data sharing" = sum(is_open_data == TRUE),
                  "Code sharing" = sum(is_open_code == TRUE)) %>%
        t() %>%
        as.data.frame() %>%
        rownames_to_column(var = "indicator") %>%
        mutate(percentage = round(V1/nrow(unique_transparency)*100, 1))


indicator_by_year =
        unique_transparency %>%
        select(pubYear_modified,
               is_coi_pred,
               is_fund_pred,
               is_register_pred,
               is_open_data,
               is_open_code) %>%
        gather("indicator", "value", -pubYear_modified) %>%
        count(pubYear_modified, indicator, value) %>%
        mutate(indicator = dplyr::recode(indicator,
                                          is_coi_pred = "COI disclosure",
                                          is_fund_pred = "Funding disclosure",
                                          is_register_pred = "Protocol registration",
                                          is_open_data = "Data sharing",
                                          is_open_code = "Code sharing")) %>%
        complete(indicator, value, pubYear_modified, fill = list(n = 0)) %>%
        group_by(pubYear_modified, indicator) %>%
        mutate(p = n / sum(n)) %>%
        filter(value) %>%
        ungroup()
```

```
# For reviews
proportions_reviews = unique_transparency %>%
        filter(is_review == T) %>%
        summarise("COI disclosure" = sum(is_coi_pred == TRUE),
                  "Funding disclosure" = sum(is_fund_pred == TRUE),
                  "Protocol registration" = sum(is_register_pred == TRUE),
                  "Data sharing" = sum(is_open_data == TRUE),
                  "Code sharing" = sum(is_open_code == TRUE)) %>%
        t() %>%
        as.data.frame() %>%
        rownames_to_column(var = "indicator") %>%
        mutate(percentage = round(V1/nrow(filter(unique_transparency, is_review == T))*100, [illegible]


indicator_by_year_reviews =
        unique_transparency %>%
        filter(is_review == T) %>%
        select(pubYear_modified,
               is_coi_pred,
               is_fund_pred,
               is_register_pred,
               is_open_data,
               is_open_code) %>%
        gather("indicator", "value", -pubYear_modified) %>%
        count(pubYear_modified, indicator, value) %>%
        mutate(indicator = dplyr::recode(indicator,
                                          is_coi_pred = "COI disclosure",
                                          is_fund_pred = "Funding disclosure",
                                          is_register_pred = "Protocol registration",
                                          is_open_data = "Data sharing",
                                          is_open_code = "Code sharing")) %>%
        complete(indicator, value, pubYear_modified, fill = list(n = 0)) %>%
        group_by(pubYear_modified, indicator) %>%
        mutate(p = n / sum(n)) %>%
        filter(value) %>%
        ungroup()


# For trials
proportions_trials = unique_transparency %>%
        filter(is_trial == T) %>%
        summarise("COI disclosure" = sum(is_coi_pred == TRUE),
```

```
                  "Funding disclosure" = sum(is_fund_pred == TRUE),
                  "Protocol registration" = sum(is_register_pred == TRUE),
                  "Data sharing" = sum(is_open_data == TRUE),
                  "Code sharing" = sum(is_open_code == TRUE)) %>%
        t() %>%
        as.data.frame() %>%
        rownames_to_column(var = "indicator") %>%
        mutate(percentage = round(V1/nrow(filter(unique_transparency, is_trial == T))*100, 1


indicator_by_year_trials =
        unique_transparency %>%
        filter(is_trial == T) %>%
        select(pubYear_modified,
               is_coi_pred,
               is_fund_pred,
               is_register_pred,
               is_open_data,
               is_open_code) %>%
        gather("indicator", "value", -pubYear_modified) %>%
        count(pubYear_modified, indicator, value) %>%
        mutate(indicator = dplyr::recode(indicator,
                                  is_coi_pred = "COI disclosure",
                                  is_fund_pred = "Funding disclosure",
                                  is_register_pred = "Protocol registration",
                                  is_open_data = "Data sharing",
                                  is_open_code = "Code sharing")) %>%
        complete(indicator, value, pubYear_modified, fill = list(n = 0)) %>%
        group_by(pubYear_modified, indicator) %>%
        mutate(p = n / sum(n)) %>%
        filter(value) %>%
        ungroup()
```

Now, we create plots:

```
### Figure 1A - left
p1 =  proportions %>%
  ggplot() +
  aes(
    x = reorder(indicator, V1),
    y = V1,
    fill = indicator
```

```
  ) +
  geom_col() +
  geom_text(aes(label = percentage), hjust = -0.1, size = 4) +
  coord_flip() +
  labs(title = "A - All articles",
       x = NULL,
       y = NULL) +
  theme_minimal() +
  theme(legend.position = "none",
        panel.grid.major.y = element_blank(),
        axis.text = element_text(size = 10),
        plot.title = element_text(size=14, face="bold")) +
  scale_fill_manual(values = c("red", viridis::viridis(6)))

### Figure 1A - right

data_ends = indicator_by_year %>%
        filter(pubYear_modified == 2024)

plasma_pal <- c("blue", viridis::plasma(n = 5))

p2 = indicator_by_year %>%
  ggplot() +
  aes(
    x = pubYear_modified,
    y = p,
    group = indicator,
    color = indicator
  ) +
  geom_line() +
  labs(title = NULL,
       y = NULL,
       x = NULL) +
   geom_text_repel(
    aes(label = indicator),
    data = data_ends,
    nudge_x = 2,
    size = 3
  ) +
  scale_y_continuous(limits = c(0, 1), labels = scales::percent) +
  theme_minimal() +
  theme(legend.position = "none") +
```

```
  scale_color_manual(values = c("red", viridis::viridis(6)))+
         theme(axis.text.x = element_text(angle = 45),
        axis.text = element_text(size = 10))


### Fig 1A
figure1A = ggarrange(p1, p2,
                    ncol = 2, nrow = 1,
                    align = "hv", common.legend = F)
```

```
Warning in grid.Call(C_textBounds, as.graphicsAnnot(x$label), x$x, x$y, :
conversion failure on '2000-2004' in 'mbcsToSbcs': dot substituted for <e2>

Warning in grid.Call(C_textBounds, as.graphicsAnnot(x$label), x$x, x$y, :
conversion failure on '2000-2004' in 'mbcsToSbcs': dot substituted for <80>

Warning in grid.Call(C_textBounds, as.graphicsAnnot(x$label), x$x, x$y, :
conversion failure on '2000-2004' in 'mbcsToSbcs': dot substituted for <93>

Warning in grid.Call(C_textBounds, as.graphicsAnnot(x$label), x$x, x$y, :
conversion failure on '2005-2009' in 'mbcsToSbcs': dot substituted for <e2>

Warning in grid.Call(C_textBounds, as.graphicsAnnot(x$label), x$x, x$y, :
conversion failure on '2005-2009' in 'mbcsToSbcs': dot substituted for <80>

Warning in grid.Call(C_textBounds, as.graphicsAnnot(x$label), x$x, x$y, :
conversion failure on '2005-2009' in 'mbcsToSbcs': dot substituted for <93>
```

```
### Figure 1B - left
p1_reviews =  proportions_reviews %>%
  ggplot() +
  aes(
    x = reorder(indicator, V1),
    y = V1,
    fill = indicator
  ) +
  geom_col() +
  geom_text(aes(label = percentage), hjust = -0.1, size = 4) +
  coord_flip() +
  labs(title = "B - Reviews",
```

```
      x = NULL,
      y = NULL) +
  theme_minimal() +
  theme(legend.position = "none",
        panel.grid.major.y = element_blank(),
        axis.text = element_text(size = 10),
        plot.title = element_text(size=14, face="bold")) +
  scale_fill_manual(values = c("red", viridis::viridis(6)))

### Figure 1B - right

data_ends_reviews = indicator_by_year_reviews %>%
        filter(pubYear_modified == 2024)

p2_reviews = indicator_by_year_reviews %>%
  ggplot() +
  aes(
    x = pubYear_modified,
    y = p,
    group = indicator,
    color = indicator
  ) +
  geom_line() +
  labs(title = NULL,
      y = "Proportion of articles",
      x = NULL) +
   geom_text_repel(
    aes(label = indicator),
    data = data_ends_reviews,
    nudge_x = 2,
    size = 3
  ) +
  scale_y_continuous(limits = c(0, 1), labels = scales::percent) +
  theme_minimal() +
  theme(legend.position = "none") +
  scale_color_manual(values = c("red", viridis::viridis(6)))+
         theme(axis.text.x = element_text(angle = 45),
        axis.text = element_text(size = 10))


### Fig 1B
figure1B = ggarrange(p1_reviews, p2_reviews,
```

```
                    ncol = 2, nrow = 1,
                    align = "hv", common.legend = F)
```

```
Warning in grid.Call(C_textBounds, as.graphicsAnnot(x$label), x$x, x$y, :
conversion failure on '2000-2004' in 'mbcsToSbcs': dot substituted for <e2>

Warning in grid.Call(C_textBounds, as.graphicsAnnot(x$label), x$x, x$y, :
conversion failure on '2000-2004' in 'mbcsToSbcs': dot substituted for <80>

Warning in grid.Call(C_textBounds, as.graphicsAnnot(x$label), x$x, x$y, :
conversion failure on '2000-2004' in 'mbcsToSbcs': dot substituted for <93>

Warning in grid.Call(C_textBounds, as.graphicsAnnot(x$label), x$x, x$y, :
conversion failure on '2005-2009' in 'mbcsToSbcs': dot substituted for <e2>

Warning in grid.Call(C_textBounds, as.graphicsAnnot(x$label), x$x, x$y, :
conversion failure on '2005-2009' in 'mbcsToSbcs': dot substituted for <80>

Warning in grid.Call(C_textBounds, as.graphicsAnnot(x$label), x$x, x$y, :
conversion failure on '2005-2009' in 'mbcsToSbcs': dot substituted for <93>
```

```
### Figure 1C - left
p1_trials =  proportions_trials %>%
  ggplot() +
  aes(
    x = reorder(indicator, V1),
    y = V1,
    fill = indicator
  ) +
  geom_col() +
  geom_text(aes(label = percentage), hjust = -0.1, size = 4) +
  coord_flip() +
  labs(title = "C - Trials",
       x = NULL,
       y = "Number of articles") +
  theme_minimal() +
  theme(legend.position = "none",
        panel.grid.major.y = element_blank(),
        axis.text = element_text(size = 10),
        plot.title = element_text(size=14, face="bold")) +
```

```
  scale_fill_manual(values = c("red", viridis::viridis(6)))

### Figure 1C - right

data_ends_trials = indicator_by_year_trials %>%
        filter(pubYear_modified == 2024)

p2_trials = indicator_by_year_trials %>%
  ggplot() +
  aes(
    x = pubYear_modified,
    y = p,
    group = indicator,
    color = indicator
  ) +
  geom_line() +
  labs(title = NULL,
       y = NULL,
       x = "Year") +
   geom_text_repel(
    aes(label = indicator),
    data = data_ends_trials,
    nudge_x = 2,
    size = 3
  ) +
  scale_y_continuous(limits = c(0, 1), labels = scales::percent) +
  theme_minimal() +
  theme(legend.position = "none") +
  scale_color_manual(values = c("red", viridis::viridis(6)))+
         theme(axis.text.x = element_text(angle = 45),
        axis.text = element_text(size = 10))


### Fig 1C
figure1C = ggarrange(p1_trials, p2_trials,
                    ncol = 2, nrow = 1,
                    align = "hv", common.legend = F)
```

```
Warning in grid.Call(C_textBounds, as.graphicsAnnot(x$label), x$x, x$y, :
conversion failure on '2000-2004' in 'mbcsToSbcs': dot substituted for <e2>

Warning in grid.Call(C_textBounds, as.graphicsAnnot(x$label), x$x, x$y, :
```

```
conversion failure on '2000-2004' in 'mbcsToSbcs': dot substituted for <80>

Warning in grid.Call(C_textBounds, as.graphicsAnnot(x$label), x$x, x$y, :
conversion failure on '2000-2004' in 'mbcsToSbcs': dot substituted for <93>

Warning in grid.Call(C_textBounds, as.graphicsAnnot(x$label), x$x, x$y, :
conversion failure on '2005-2009' in 'mbcsToSbcs': dot substituted for <e2>

Warning in grid.Call(C_textBounds, as.graphicsAnnot(x$label), x$x, x$y, :
conversion failure on '2005-2009' in 'mbcsToSbcs': dot substituted for <80>

Warning in grid.Call(C_textBounds, as.graphicsAnnot(x$label), x$x, x$y, :
conversion failure on '2005-2009' in 'mbcsToSbcs': dot substituted for <93>
```

```r
# Figure 1
figure1 = ggarrange(figure1A, figure1B, figure1C,
                    ncol = 1, nrow = 3,
                    align = "hv", common.legend = F)


#ggsave("figures/Figure1.tiff", figure1, width = 35, height = 30, units = "cm", dpi = 800, c
#ggsave("figures/Figure1.png", figure1, width = 35, height = 30, units = "cm", dpi = 800)
```

```r
figure1
```

```
Warning in grid.Call.graphics(C_text, as.graphicsAnnot(x$label), x$x, x$y, :
conversion failure on '2000-2004' in 'mbcsToSbcs': dot substituted for <e2>

Warning in grid.Call.graphics(C_text, as.graphicsAnnot(x$label), x$x, x$y, :
conversion failure on '2000-2004' in 'mbcsToSbcs': dot substituted for <80>

Warning in grid.Call.graphics(C_text, as.graphicsAnnot(x$label), x$x, x$y, :
conversion failure on '2000-2004' in 'mbcsToSbcs': dot substituted for <93>

Warning in grid.Call.graphics(C_text, as.graphicsAnnot(x$label), x$x, x$y, :
conversion failure on '2005-2009' in 'mbcsToSbcs': dot substituted for <e2>

Warning in grid.Call.graphics(C_text, as.graphicsAnnot(x$label), x$x, x$y, :
conversion failure on '2005-2009' in 'mbcsToSbcs': dot substituted for <80>
```

```
Warning in grid.Call.graphics(C_text, as.graphicsAnnot(x$label), x$x, x$y, :
conversion failure on '2005-2009' in 'mbcsToSbcs': dot substituted for <93>

Warning in grid.Call.graphics(C_text, as.graphicsAnnot(x$label), x$x, x$y, :
conversion failure on '2000-2004' in 'mbcsToSbcs': dot substituted for <e2>

Warning in grid.Call.graphics(C_text, as.graphicsAnnot(x$label), x$x, x$y, :
conversion failure on '2000-2004' in 'mbcsToSbcs': dot substituted for <80>

Warning in grid.Call.graphics(C_text, as.graphicsAnnot(x$label), x$x, x$y, :
conversion failure on '2000-2004' in 'mbcsToSbcs': dot substituted for <93>

Warning in grid.Call.graphics(C_text, as.graphicsAnnot(x$label), x$x, x$y, :
conversion failure on '2005-2009' in 'mbcsToSbcs': dot substituted for <e2>

Warning in grid.Call.graphics(C_text, as.graphicsAnnot(x$label), x$x, x$y, :
conversion failure on '2005-2009' in 'mbcsToSbcs': dot substituted for <80>

Warning in grid.Call.graphics(C_text, as.graphicsAnnot(x$label), x$x, x$y, :
conversion failure on '2005-2009' in 'mbcsToSbcs': dot substituted for <93>

Warning in grid.Call.graphics(C_text, as.graphicsAnnot(x$label), x$x, x$y, :
conversion failure on '2000-2004' in 'mbcsToSbcs': dot substituted for <e2>

Warning in grid.Call.graphics(C_text, as.graphicsAnnot(x$label), x$x, x$y, :
conversion failure on '2000-2004' in 'mbcsToSbcs': dot substituted for <80>

Warning in grid.Call.graphics(C_text, as.graphicsAnnot(x$label), x$x, x$y, :
conversion failure on '2000-2004' in 'mbcsToSbcs': dot substituted for <93>

Warning in grid.Call.graphics(C_text, as.graphicsAnnot(x$label), x$x, x$y, :
conversion failure on '2005-2009' in 'mbcsToSbcs': dot substituted for <e2>

Warning in grid.Call.graphics(C_text, as.graphicsAnnot(x$label), x$x, x$y, :
conversion failure on '2005-2009' in 'mbcsToSbcs': dot substituted for <80>

Warning in grid.Call.graphics(C_text, as.graphicsAnnot(x$label), x$x, x$y, :
conversion failure on '2005-2009' in 'mbcsToSbcs': dot substituted for <93>
```

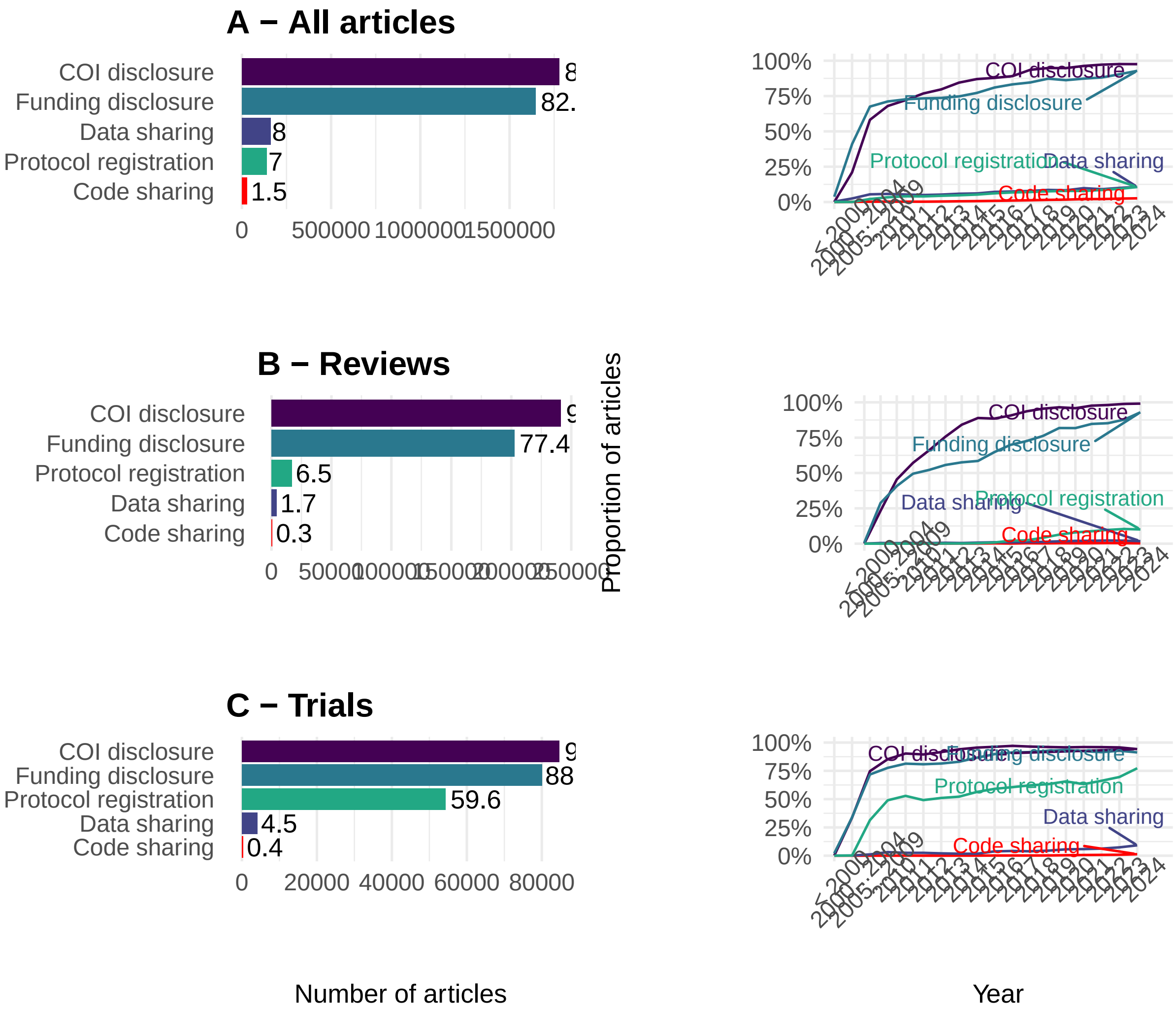


Now, testing the correlation between year and each transparency indicator:

```
indicator_by_year_all =
        unique_transparency %>%
        select(year_firstpub,
               is_coi_pred,
               is_fund_pred,
               is_register_pred,
               is_open_data,
               is_open_code) %>%
        gather("indicator", "value", -year_firstpub) %>%
        count(year_firstpub, indicator, value) %>%
        mutate(indicator = dplyr::recode(indicator,
                                         is_coi_pred = "COI disclosure",
```

```
                                  is_fund_pred = "Funding disclosure",
                                  is_register_pred = "Protocol registration",
                                  is_open_data = "Data sharing",
                                  is_open_code = "Code sharing")) %>%
      complete(indicator, value, year_firstpub, fill = list(n = 0)) %>%
      group_by(year_firstpub, indicator) %>%
      mutate(p = n / sum(n)) %>%
      filter(value) %>%
      ungroup()

cor.test(filter(indicator_by_year_all, indicator == "COI disclosure")$year_firstpub, filter(
```

```

    Pearson's product-moment correlation

data:  filter(indicator_by_year_all, indicator == "COI disclosure")$year_firstpub and filter
t = 9.7513, df = 121, p-value < 2.2e-16
alternative hypothesis: true correlation is not equal to 0
95 percent confidence interval:
 0.5510357 0.7520713
sample estimates:
     cor
0.663358
```

```
cor.test(filter(indicator_by_year_all, indicator == "Funding disclosure")$year_firstpub, fil
```

```

    Pearson's product-moment correlation

data:  filter(indicator_by_year_all, indicator == "Funding disclosure")$year_firstpub and fil
t = 11.306, df = 121, p-value < 2.2e-16
alternative hypothesis: true correlation is not equal to 0
95 percent confidence interval:
 0.6181589 0.7931467
sample estimates:
      cor
0.7167549
```

```
cor.test(filter(indicator_by_year_all, indicator == "Protocol registration")$year_firstpub, 
```

```
    Pearson's product-moment correlation

data:  filter(indicator_by_year_all, indicator == "Protocol registration")$year_firstpub and
t = 7.9115, df = 121, p-value = 1.355e-12
alternative hypothesis: true correlation is not equal to 0
95 percent confidence interval:
 0.4537639 0.6896400
sample estimates:
      cor
0.5838931
```

```
cor.test(filter(indicator_by_year_all, indicator == "Data sharing")$year_firstpub, filter(inc
```

```
    Pearson's product-moment correlation

data:  filter(indicator_by_year_all, indicator == "Data sharing")$year_firstpub and filter(in
t = 10.47, df = 121, p-value < 2.2e-16
alternative hypothesis: true correlation is not equal to 0
95 percent confidence interval:
 0.5836425 0.7722217
sample estimates:
      cor
0.6894406
```

```
cor.test(filter(indicator_by_year_all, indicator == "Code sharing")$year_firstpub, filter(inc
```

```
    Pearson's product-moment correlation

data:  filter(indicator_by_year_all, indicator == "Code sharing")$year_firstpub and filter(in
t = 6.6146, df = 121, p-value = 1.068e-09
alternative hypothesis: true correlation is not equal to 0
95 percent confidence interval:
 0.3722595 0.6344817
sample estimates:
     cor
0.515332
```

##### 2.5.2 Appendix 4: Yearly trends for each field

```
indicator_by_year_all_fields =
        transparency %>%
        select(category,
                pubYear_modified,
               is_coi_pred,
               is_fund_pred,
               is_register_pred,
               is_open_data,
               is_open_code) %>%
        gather("indicator", "value", -pubYear_modified, -category) %>%
        count(category, pubYear_modified, indicator, value) %>%
        mutate(indicator = dplyr::recode(indicator,
                                          is_coi_pred = "COI Disclosure",
                                          is_fund_pred = "Funding disclosure",
                                          is_register_pred = "Protocol registration",
                                          is_open_data = "Data sharing",
                                          is_open_code = "Code sharing")) %>%
        complete(indicator, value, pubYear_modified, category, fill = list(n = 0)) %>%
        group_by(category, pubYear_modified, indicator) %>%
        mutate(p = n / sum(n)) %>%
        filter(value) %>%
        ungroup()

transparency_all_fields = indicator_by_year_all_fields %>%
        ggplot() +
        aes(x = pubYear_modified,
            y = p,
            group = indicator,
            color = indicator) +
        geom_line(size = 0.75) +
        facet_wrap(~ category, ncol = 10) +
        scale_y_continuous(limits = c(0, 1),
                           labels = scales::percent) +
        scale_color_discrete(name = NULL) +
        scale_fill_discrete(breaks = c("COI Disclosure",
                                       "Funding disclosure",
                                       "Protocol registration",
                                       "Data sharing",
                                       "Code sharing")) +
        labs(y = "Proportion of articles\n",
```

```
            x = "\nYear") +
        theme(panel.grid.minor = element_blank(),
              legend.position = c(0.955, 0.001),
              axis.text.x = element_text(angle = 70, vjust = 0.9, hjust=1, size = 7))

transparency_all_fields
```

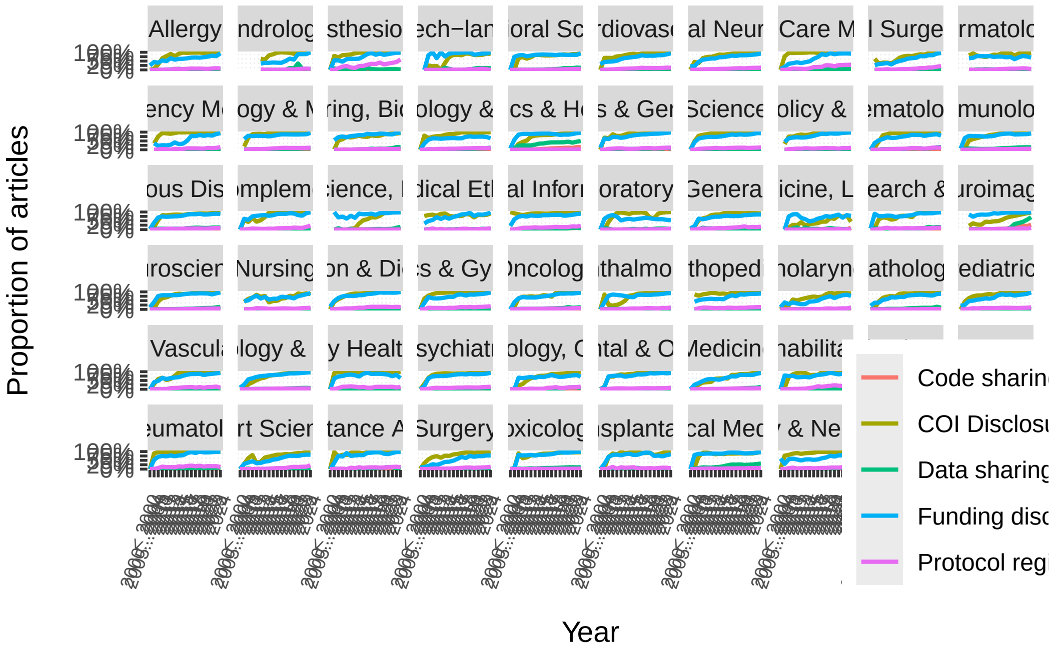


```
#ggsave("appendix/Appendix4.tiff", transparency_all_fields, width = 18, height = 8, units = "
#ggsave("appendix/Appendix4.png", transparency_all_fields, width = 18, height = 8, units = "i
```

###### 2.5.2.1 GLMM: Yearly trend of adherence to transparency practices among different fields

COI disclosure:

```
coi_glmer = glmer(is_coi_pred ~ year_firstpub + (1|category), data = transparency, family = b

summary(coi_glmer)
```

```
Generalized linear mixed model fit by maximum likelihood (Adaptive
  Gauss-Hermite Quadrature, nAGQ = 10) [glmerMod]
 Family: binomial  ( logit )
```

```
Formula: is_coi_pred ~ year_firstpub + (1 | category)
   Data: transparency
Control: glmerControl(optimizer = "bobyqa", optCtrl = list(maxfun = 2e+05))

     AIC      BIC   logLik deviance df.resid
 1227570  1227608  -613782  1227564  2471010

Scaled residuals:
     Min       1Q   Median       3Q      Max
-25.3150   0.1452   0.1979   0.2973  28.9552

Random effects:
 Groups   Name        Variance Std.Dev.
 category (Intercept) 0.8      0.8944
Number of obs: 2471013, groups:  category, 59

Fixed effects:
                Estimate Std. Error z value Pr(>|z|)
(Intercept)   -4.818e+02  1.950e-01   -2471   <2e-16 ***
year_firstpub  2.401e-01  1.113e-04    2157   <2e-16 ***
---
Signif. codes:  0 '***' 0.001 '**' 0.01 '*' 0.05 '.' 0.1 ' ' 1

Correlation of Fixed Effects:
            (Intr)
year_frstpb -0.857
optimizer (bobyqa) convergence code: 0 (OK)
Model failed to converge with max|grad| = 0.0359188 (tol = 0.002, component 1)
Model is nearly unidentifiable: very large eigenvalue
 - Rescale variables?
Model is nearly unidentifiable: large eigenvalue ratio
 - Rescale variables?
```

Funding disclosure:

```
fund_glmer = glmer(is_fund_pred ~ year_firstpub + (1|category), data = transparency, family =

summary(fund_glmer)
```

```
Generalized linear mixed model fit by maximum likelihood (Adaptive
  Gauss-Hermite Quadrature, nAGQ = 10) [glmerMod]
 Family: binomial  ( logit )
```

```
Formula: is_fund_pred ~ year_firstpub + (1 | category)
   Data: transparency
Control: glmerControl(optimizer = "bobyqa", optCtrl = list(maxfun = 2e+05))

     AIC      BIC   logLik deviance df.resid 
 2012165  2012204 -1006080  2012159  2471010 

Scaled residuals: 
   Min     1Q Median     3Q    Max 
-6.220  0.267  0.346  0.450 46.638 

Random effects:
 Groups   Name        Variance Std.Dev.
 category (Intercept) 0.3603   0.6003  
Number of obs: 2471013, groups:  category, 59

Fixed effects:
                Estimate Std. Error z value Pr(>|z|)    
(Intercept)   -2.651e+02  2.815e-01  -941.6   <2e-16 ***
year_firstpub  1.321e-01  1.428e-04   925.1   <2e-16 ***
---
Signif. codes:  0 '***' 0.001 '**' 0.01 '*' 0.05 '.' 0.1 ' ' 1

Correlation of Fixed Effects:
            (Intr)
year_frstpb -0.963
optimizer (bobyqa) convergence code: 0 (OK)
Model failed to converge with max|grad| = 0.0308105 (tol = 0.002, component 1)
Model is nearly unidentifiable: very large eigenvalue
 - Rescale variables?
Model is nearly unidentifiable: large eigenvalue ratio
 - Rescale variables?
```

Protocol registration:

```r
register_glmer = glmer(is_register_pred ~ year_firstpub + (1|category), data = transparency,

summary(register_glmer)
```

```
Generalized linear mixed model fit by maximum likelihood (Adaptive
  Gauss-Hermite Quadrature, nAGQ = 10) [glmerMod]
 Family: binomial  ( logit )
```

```
Formula: is_register_pred ~ year_firstpub + (1 | category)
   Data: transparency
Control: glmerControl(optimizer = "bobyqa", optCtrl = list(maxfun = 2e+05))

     AIC      BIC   logLik deviance df.resid
 1173022  1173060  -586508  1173016  2471010

Scaled residuals:
    Min      1Q  Median      3Q     Max
-0.8765 -0.3208 -0.2424 -0.1753 18.4747

Random effects:
 Groups   Name        Variance Std.Dev.
 category (Intercept) 0.7428   0.8619
Number of obs: 2471013, groups:  category, 59

Fixed effects:
                Estimate Std. Error z value Pr(>|z|)
(Intercept)   -1.717e+02  1.917e-01  -895.9   <2e-16 ***
year_firstpub  8.370e-02  1.095e-04   764.6   <2e-16 ***
---
Signif. codes:  0 '***' 0.001 '**' 0.01 '*' 0.05 '.' 0.1 ' ' 1

Correlation of Fixed Effects:
            (Intr)
year_frstpb -0.862
optimizer (bobyqa) convergence code: 0 (OK)
Model failed to converge with max|grad| = 0.00453864 (tol = 0.002, component 1)
Model is nearly unidentifiable: very large eigenvalue
 - Rescale variables?
Model is nearly unidentifiable: large eigenvalue ratio
 - Rescale variables?
```

Data sharing:

```r
data_glmer = glmer(is_open_data ~ year_firstpub + (1|category), data = transparency, family =

summary(data_glmer)
```

```
Generalized linear mixed model fit by maximum likelihood (Adaptive
  Gauss-Hermite Quadrature, nAGQ = 10) [glmerMod]
 Family: binomial  ( logit )
```

```
Formula: is_open_data ~ year_firstpub + (1 | category)
   Data: transparency
Control: glmerControl(optimizer = "bobyqa", optCtrl = list(maxfun = 2e+05))

      AIC       BIC    logLik  deviance  df.resid
1148882.8 1148920.9 -574438.4 1148876.8   2471010

Scaled residuals:
    Min      1Q  Median      3Q     Max
-1.0861 -0.2812 -0.2184 -0.1649 18.5164

Random effects:
 Groups   Name        Variance Std.Dev.
 category (Intercept) 0.5648   0.7515
Number of obs: 2471013, groups:  category, 59

Fixed effects:
                Estimate Std. Error z value Pr(>|z|)
(Intercept)   -2.087e+02  2.145e-01  -973.2   <2e-16 ***
year_firstpub  1.019e-01  1.162e-04   876.7   <2e-16 ***
---
Signif. codes:  0 '***' 0.001 '**' 0.01 '*' 0.05 '.' 0.1 ' ' 1

Correlation of Fixed Effects:
            (Intr)
year_frstpb -0.909
optimizer (bobyqa) convergence code: 0 (OK)
Model failed to converge with max|grad| = 0.00329572 (tol = 0.002, component 1)
Model is nearly unidentifiable: very large eigenvalue
 - Rescale variables?
Model is nearly unidentifiable: large eigenvalue ratio
 - Rescale variables?
```

Code sharing:

```r
code_glmer = glmer(is_open_code ~ year_firstpub + (1|category), data = transparency, family =

summary(code_glmer)
```

```
Generalized linear mixed model fit by maximum likelihood (Adaptive
  Gauss-Hermite Quadrature, nAGQ = 10) [glmerMod]
 Family: binomial  ( logit )
```

```
Formula: is_open_code ~ year_firstpub + (1 | category)
   Data: transparency
Control: glmerControl(optimizer = "bobyqa", optCtrl = list(maxfun = 2e+05))

      AIC       BIC    logLik  deviance  df.resid
 322897.9  322936.1 -161446.0  322891.9   2471010

Scaled residuals:
   Min     1Q Median     3Q    Max
-0.582 -0.120 -0.087 -0.055 78.146

Random effects:
 Groups   Name        Variance Std.Dev.
 category (Intercept) 1.311    1.145
Number of obs: 2471013, groups:  category, 59

Fixed effects:
                Estimate Std. Error z value Pr(>|z|)
(Intercept)   -4.206e+02  4.208e-01  -999.5   <2e-16 ***
year_firstpub  2.057e-01  2.210e-04   930.9   <2e-16 ***
---
Signif. codes:  0 '***' 0.001 '**' 0.01 '*' 0.05 '.' 0.1 ' ' 1

Correlation of Fixed Effects:
            (Intr)
year_frstpb -0.942
optimizer (bobyqa) convergence code: 0 (OK)
Model failed to converge with max|grad| = 0.0226471 (tol = 0.002, component 1)
Model is nearly unidentifiable: very large eigenvalue
 - Rescale variables?
Model is nearly unidentifiable: large eigenvalue ratio
 - Rescale variables?
```

##### 2.5.3 Appendix 3

```
percent_calc = function(indicator){
        x = round(sum(indicator)/n()*100, 1)
        return(x)
}

transparency_by_field = transparency %>%
```

```
        group_by(category) %>%
        summarise("COI Disclosure" = sum(is_coi_pred == TRUE),
                  "COI Disclosure %" = percent_calc(is_coi_pred == TRUE),
                  "Funding disclosure" = sum(is_fund_pred == TRUE),
                  "Funding disclosure %" = percent_calc(is_fund_pred == TRUE),
                  "Protocol registration" = sum(is_register_pred == TRUE),
                  "Protocol registration %" = percent_calc(is_register_pred == TRUE),
                  "Data sharing" = sum(is_open_data == TRUE),
                  "Data sharing %" = percent_calc(is_open_data == TRUE),
                  "Code sharing" = sum(is_open_code == TRUE),
                  "Code sharing %" = percent_calc(is_open_code == TRUE))

# write.csv(transparency_by_field, "appendix/Appendix3.csv", row.names = F)
```

Now, min and max for each indicator based on category. For COI disclosure:

```
transparency_by_field %>% arrange(desc(`COI Disclosure %`)) %>% filter(row_number() %in% c(1
```

```
# A tibble: 6 x 2
  category             `COI Disclosure %`
  <chr>                             <dbl>
1 Rheumatology                       98.1
2 Primary Health Care                97.4
3 Emergency Medicine                 96.8
4 Neuroimaging                       71.5
5 Toxicology                         67.5
6 Medicine, Legal                    61.6
```

For funding disclosure:

```
transparency_by_field %>% arrange(desc(`Funding disclosure %`)) %>% filter(row_number() %in%
```

```
# A tibble: 6 x 2
  category                                   `Funding disclosure %`
  <chr>                                                       <dbl>
1 Neuroimaging                                                 94.9
2 Materials Science, Biomaterials                              94
3 Audiology & Speech-language Pathology                        93.3
4 Critical Care Medicine                                       64.8
5 Andrology                                                    64.7
6 Medical Laboratory Technology                                60.3
```

Protocol registration:

```
transparency_by_field %>% arrange(desc(`Protocol registration %`)) %>% filter(row_number() %
```

```
# A tibble: 6 x 2
  category                          `Protocol registration %`
  <chr>                                                 <dbl>
1 Anesthesiology                                         34.5
2 Rehabilitation                                         17.2
3 Critical Care Medicine                                 15.2
4 Virology                                                1.2
5 Genetics & Heredity                                     0.9
6 Materials Science, Biomaterials                         0.3
```

Data sharing:

```
transparency_by_field %>% arrange(desc(`Data sharing %`)) %>% filter(row_number() %in% c(1:3
```

```
# A tibble: 6 x 2
  category            `Data sharing %`
  <chr>                          <dbl>
1 Genetics & Heredity             37.9
2 Neuroimaging                    24.7
3 Virology                        23.5
4 Orthopedics                      1.6
5 Primary Health Care              1.6
6 Surgery                          1.6
```

Code sharing:

```
transparency_by_field %>% arrange(desc(`Code sharing %`)) %>% filter(row_number() %in% c(1:3
```

```
# A tibble: 6 x 2
  category                                 `Code sharing %`
  <chr>                                               <dbl>
1 Neuroimaging                                         12.4
2 Genetics & Heredity                                   7.6
3 Medical Informatics                                   6.3
4 Integrative & Complementary Medicine                  0.1
5 Nursing                                               0.1
6 Orthopedics                                           0.1
```

#### 2.6 Association with impact factor and number of citations

Adding impact factor variable:

```
impact_factors = read.csv("data/journals.csv")

matches = str_extract(unique_transparency$journalIssn, paste(impact_factors$ISSN, collapse =
matching_rows = !is.na(matches)

unique_transparency$jif2020[matching_rows] = impact_factors$X2020.JIF[match(matches[matching_

unique_transparency$jif2020 = as.numeric(unique_transparency$jif2020)
```

```
Warning: NAs introduced by coercion
```

##### 2.6.1 Figure 2

```
unique_transparency$jif2020_5 = factor(quant_groups(unique_transparency$jif2020, 5))
```

```
Observations per group: 400075, 401982, 397605, 411196, 388541. 3556 missing.
```

```
levels(unique_transparency$jif2020_5) = c("Q1", "Q2", "Q3", "Q4", "Q5")
unique_transparency = apply_labels(unique_transparency,
                        jif2020_5 = "Journal Impact Factor",
                        is_coi_pred = "COI disclosure",
                        is_fund_pred = "Funding disclosure",
                        is_register_pred = "Protocol registeration",
                        is_open_data = "Data sharing",
                        is_open_code = "Code sharing")



coi_jif = glm(is_coi_pred ~ jif2020_5 + year_firstpub, family = "binomial", data = unique_tra
fund_jif = glm(is_fund_pred ~ jif2020_5 + year_firstpub, family = "binomial", data = unique_t
reg_jif = glm(is_register_pred ~ jif2020_5 + year_firstpub, family = "binomial", data = uniqu
data_jif = glm(is_open_data ~ jif2020_5 + year_firstpub, family = "binomial", data = unique_t
code_jif = glm(is_open_code ~ jif2020_5 + year_firstpub, family = "binomial", data = unique_t
```

```
terms = paste(as.character(unique(unique_transparency$year_firstpub)), collapse = ", ")

jif_plot = plot_models(coi_jif, fund_jif, reg_jif, data_jif, code_jif, rm.terms = "year_firs

unique_transparency$citedByCount_4 = factor(ifelse(unique_transparency$citedByCount == 0,"0"
                                        ifelse(unique_transparency$citedByCount >0 & unique_
                                               ifelse(unique_transparency$citedByCount >10 &
                                                      ifelse(unique_transparency$citedByCoun
                                   c("0","1-10","11-100","> 100"))

coi_cite = glm(is_coi_pred ~ citedByCount_4 + year_firstpub, family = "binomial", data = uniq
fund_cite = glm(is_fund_pred ~ citedByCount_4 + year_firstpub, family = "binomial", data = un
reg_cite = glm(is_register_pred ~ citedByCount_4 + year_firstpub, family = "binomial", data =
data_cite = glm(is_open_data ~ citedByCount_4 + year_firstpub, family = "binomial", data = un
code_cite = glm(is_open_code ~ citedByCount_4 + year_firstpub, family = "binomial", data = un

cite_plot = plot_models(coi_cite, fund_cite, reg_cite, data_cite, code_cite, rm.terms = "yea


figure2A = ggarrange(jif_plot + theme(legend.title=element_blank()), cite_plot + theme(legenc


# Reviews
reviews = unique_transparency %>% filter(is_review == T)

coi_jif_reviews = glm(is_coi_pred ~ jif2020_5 + year_firstpub, family = "binomial", data = re
fund_jif_reviews = glm(is_fund_pred ~ jif2020_5 + year_firstpub, family = "binomial", data =
reg_jif_reviews = glm(is_register_pred ~ jif2020_5 + year_firstpub, family = "binomial", data
data_jif_reviews = glm(is_open_data ~ jif2020_5 + year_firstpub, family = "binomial", data =
code_jif_reviews = glm(is_open_code ~ jif2020_5 + year_firstpub, family = "binomial", data =

jif_plot_reviews = plot_models(coi_jif_reviews, fund_jif_reviews, reg_jif_reviews, data_jif_r

coi_cite_reviews = glm(is_coi_pred ~ citedByCount_4 + year_firstpub, family = "binomial", dat
fund_cite_reviews = glm(is_fund_pred ~ citedByCount_4 + year_firstpub, family = "binomial", c
reg_cite_reviews = glm(is_register_pred ~ citedByCount_4 + year_firstpub, family = "binomial"
data_cite_reviews = glm(is_open_data ~ citedByCount_4 + year_firstpub, family = "binomial", c
code_cite_reviews = glm(is_open_code ~ citedByCount_4 + year_firstpub, family = "binomial", c

cite_plot_reviews = plot_models(coi_cite_reviews, fund_cite_reviews, reg_cite_reviews, data_c

figure2B = ggarrange(jif_plot_reviews + theme(legend.title=element_blank()), cite_plot_review
```

```
# Trials
trials = unique_transparency %>% filter(is_trial == T)

coi_jif_trials = glm(is_coi_pred ~ jif2020_5 + year_firstpub, family = "binomial", data = tri
fund_jif_trials = glm(is_fund_pred ~ jif2020_5 + year_firstpub, family = "binomial", data = t
reg_jif_trials = glm(is_register_pred ~ jif2020_5 + year_firstpub, family = "binomial", data
data_jif_trials = glm(is_open_data ~ jif2020_5 + year_firstpub, family = "binomial", data = t
code_jif_trials = glm(is_open_code ~ jif2020_5 + year_firstpub, family = "binomial", data = t

jif_plot_trials = plot_models(coi_jif_trials, fund_jif_trials, reg_jif_trials, data_jif_trial

coi_cite_trials = glm(is_coi_pred ~ citedByCount_4 + year_firstpub, family = "binomial", data
fund_cite_trials = glm(is_fund_pred ~ citedByCount_4 + year_firstpub, family = "binomial", da
reg_cite_trials = glm(is_register_pred ~ citedByCount_4 + year_firstpub, family = "binomial"
data_cite_trials = glm(is_open_data ~ citedByCount_4 + year_firstpub, family = "binomial", da
code_cite_trials = glm(is_open_code ~ citedByCount_4 + year_firstpub, family = "binomial", da

cite_plot_trials = plot_models(coi_cite_trials, fund_cite_trials, reg_cite_trials, data_cite_


figure2C = ggarrange(jif_plot_trials + theme(legend.title=element_blank()), cite_plot_trials

figure2 = ggarrange(figure2A, figure2B, figure2C, ncol = 1, nrow = 3, common.legend = T, labe

# ggsave("figures/Figure2.png", figure2, dpi = 800, width = 27, height = 30, units = "cm")
# ggsave("figures/Figure2.tiff", figure2, dpi = 800, width = 27, height = 30, units = "cm", c
```

```
figure2
```

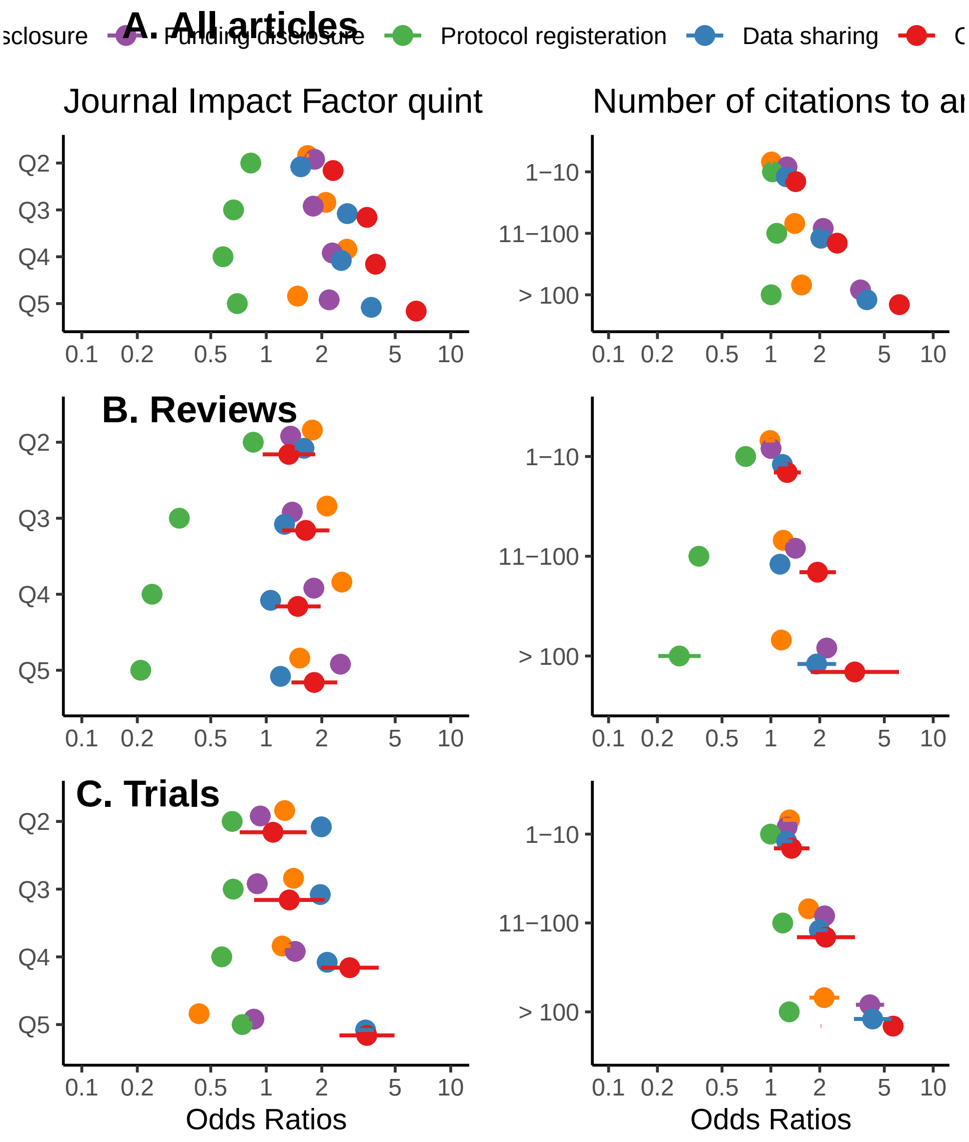


#### 2.7 Publisher differences

Let's take a look at the top most and least transparent publishers in each indicator.

##### 2.7.1 Conflict of interest disclosure

```
unique_transparency %>%
        select(is_coi_pred, scimago_publisher) %>%
```

```
        group_by(scimago_publisher) %>%
        summarise(coi = sum(is_coi_pred)/n()*100) %>%
        arrange(desc(coi)) %>%
        head(5)
```

```
# A tibble: 5 x 2
  scimago_publisher                            coi
  <chr>                                      <dbl>
1 AME Publishing Company                       100
2 Academia Nacional de Medicina                100
3 Academy of Medical Sciences of I.R. Iran     100
4 Acta Cardiologica                            100
5 American Academy of Pediatrics               100
```

It seems many publishers had 100% transparency. Let's check how many are these:

```
nrow(unique_transparency %>%
        select(is_coi_pred, scimago_publisher) %>%
        group_by(scimago_publisher) %>%
        summarise(coi = sum(is_coi_pred)/n()*100) %>%
        filter(coi == 100))
```

```
[1] 71
```

Now, the top lowest adherence:

```
unique_transparency %>%
        select(is_coi_pred, scimago_publisher) %>%
        group_by(scimago_publisher) %>%
        summarise(coi = sum(is_coi_pred)/n()*100) %>%
        arrange(coi) %>%
        head(5)
```

```
# A tibble: 5 x 2
  scimago_publisher                                coi
  <chr>                                          <dbl>
1 Histology and Histopathology                       0
2 International Union of Crystallography             0
3 National Co-ordinating Centre for HTA              0
4 Sociedad Espanola de Medicina Interna (SEMI)       0
5 Sub Rosa                                           0
```

It seems there are more than one. Let's count them:

```
nrow(unique_transparency %>%
        select(is_coi_pred, scimago_publisher) %>%
        group_by(scimago_publisher) %>%
        summarise(coi = sum(is_coi_pred)/n()*100) %>%
        filter(coi == 0))
```

```
[1] 8
```

##### 2.7.2 Funding disclosure

```
unique_transparency %>%
        select(is_fund_pred, scimago_publisher) %>%
        group_by(scimago_publisher) %>%
        summarise(fund = sum(is_fund_pred)/n()*100) %>%
        arrange(desc(fund)) %>%
        head(5)
```

```
# A tibble: 5 x 2
  scimago_publisher                                    fund
  <chr>                                               <dbl>
1 AO Research Institute                                 100
2 Alcohol Research Documentation, Inc.                  100
3 American Academy of Pediatrics                        100
4 American Association for the Advancement of Science   100
5 American Association of Immunologists                 100
```

```
unique_transparency %>%
        select(is_fund_pred, scimago_publisher) %>%
        group_by(scimago_publisher) %>%
        summarise(fund = sum(is_fund_pred)/n()*100) %>%
        arrange(fund) %>%
        head(5)
```

```
# A tibble: 5 x 2
  scimago_publisher                      fund
  <chr>                                 <dbl>
1 Acta Cardiologica                         0
```

```
2 Cairo University                          0
3 Deutscher Arzte-Verlag                    0
4 Histology and Histopathology              0
5 National Co-ordinating Centre for HTA     0
```

##### 2.7.3 Protocol registration

```
unique_transparency %>%
        select(is_register_pred, scimago_publisher) %>%
        group_by(scimago_publisher) %>%
        summarise(register = sum(is_register_pred)/n()*100) %>%
        arrange(desc(register)) %>%
        head(5)
```

```
# A tibble: 5 x 2
  scimago_publisher              register
  <chr>                             <dbl>
1 American Society for Nutrition     40.0
2 OceanSide Publications Inc.        35
3 Lancet Publishing Group            29.5
4 Massachussetts Medical Society     27.0
5 American Heart Association         25.4
```

```
unique_transparency %>%
        select(is_register_pred, scimago_publisher) %>%
        group_by(scimago_publisher) %>%
        summarise(register = sum(is_register_pred)/n()*100) %>%
        arrange(register) %>%
        head(5)
```

```
# A tibble: 5 x 2
  scimago_publisher                    register
  <chr>                                   <dbl>
1 AO Research Institute                       0
2 Academia Nacional de Medicina               0
3 Acoustical Society of America               0
4 Acta Cardiologica                           0
5 Alcohol Research Documentation, Inc.        0
```

##### 2.7.4 Data sharing

```
unique_transparency %>%
        select(is_open_data, scimago_publisher) %>%
        group_by(scimago_publisher) %>%
        summarise(data = sum(is_open_data)/n()*100) %>%
        arrange(desc(data)) %>%
        head(5)
```

```
# A tibble: 5 x 2
  scimago_publisher                                    data
  <chr>                                               <dbl>
1 International Union of Crystallography               100
2 American Association for the Advancement of Science  67.0
3 Genetics Society of America                          62.9
4 Cold Spring Harbor Laboratory Press                  60.3
5 Microbiology Society                                 58.6
```

```
unique_transparency %>%
        select(is_open_data, scimago_publisher) %>%
        group_by(scimago_publisher) %>%
        summarise(data = sum(is_open_data)/n()*100) %>%
        arrange(data) %>%
        head(5)
```

```
# A tibble: 5 x 2
  scimago_publisher                                   data
  <chr>                                              <dbl>
1 Academia Nacional de Medicina                          0
2 Acta Cardiologica                                      0
3 American Cleft Palate Craniofacial Association         0
4 American College of Allergy, Asthma and Immunology     0
5 American Dental Association                            0
```

##### 2.7.5 Code sharing

```
unique_transparency %>%
        select(is_open_code, scimago_publisher) %>%
```

```
        group_by(scimago_publisher) %>%
        summarise(code = sum(is_open_code)/n()*100) %>%
        arrange(desc(code)) %>%
        head(5)
```

```
# A tibble: 5 x 2
  scimago_publisher                       code
  <chr>                                  <dbl>
1 Cold Spring Harbor Laboratory Press  40.5
2 MIT Press Journals                     35.1
3 American College of Physicians         20
4 Thieme Medical Publishers              20
5 Microbiology Society                   18.7
```

```
unique_transparency %>%
        select(is_open_code, scimago_publisher) %>%
        group_by(scimago_publisher) %>%
        summarise(code = sum(is_open_code)/n()*100) %>%
        arrange(code) %>%
        head(5)
```

```
# A tibble: 5 x 2
  scimago_publisher                                code
  <chr>                                           <dbl>
1 ""                                                  0
2 "AO Research Institute"                             0
3 "AOSIS (Pty) Ltd"                                   0
4 "Academia Nacional de Medicina"                     0
5 "Academy of Managed Care Pharmacy (AMCP)"           0
```
