## Supplementary figures and images for "Research Transparency in 59 Disciplines of Clinical Medicine: A Meta-Research Study"

### Appendix 4

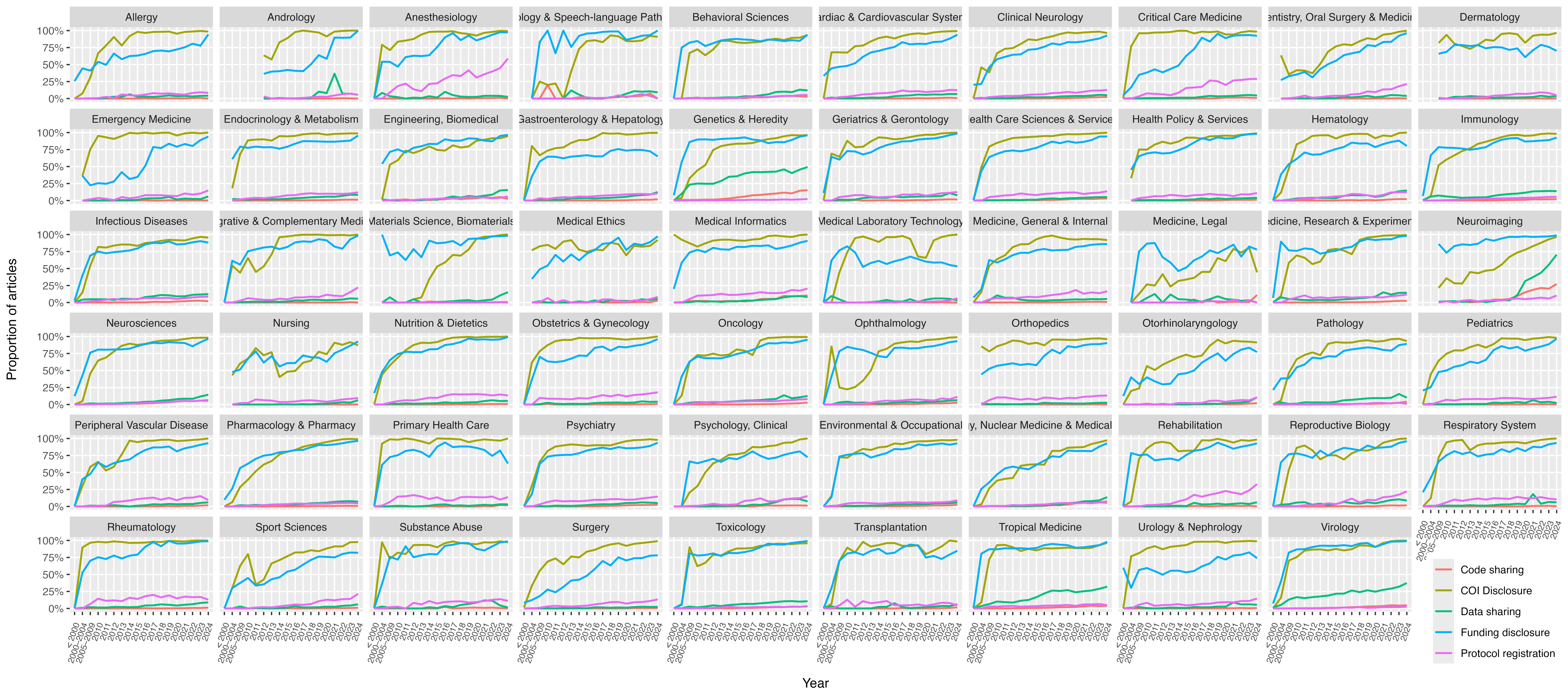
